# Mosaic variegated aneuploidy syndrome with tetraploid, and predisposition to male infertility triggered by mutant *CEP192*

**DOI:** 10.1101/2023.07.22.23292907

**Authors:** Jihong Guo, Wen-bin He, Lei Dai, Fen Tian, Zhenqing Luo, Fang Shen, Ming Tu, Yu Zheng, Liu Zhao, Chen Tan, Yongteng Guo, Lan-Lan Meng, Wei Liu, Mei Deng, Xinghan Wu, Yu Peng, Shuju Zhang, Guang-Xiu Lu, Ge Lin, Hua Wang, Yue-Qiu Tan, Yongjia Yang

## Abstract

In the present study, we report on mosaic variegated aneuploidy (MVA) syndrome with tetraploidy and predisposition to infertility in a family. Sequencing analysis identified that the *CEP192* biallelic variants (c.1912C>T/p.H638Y and c.5750A>G/p.N1917S) segregated with microcephaly, short stature, limb–extremity dysplasia, and reduced testicular size, while *CEP192* monoallelic variants segregated with infertility and/or reduced testicular size in the family. In 1,264 unrelated patients, variant screening for *CEP192* identified a same variant (c.5750A>G/p.N1917S) and other variants significantly associated with infertility. Two lines of *Cep192* mice model that are equivalent to human variants were generated. Embryos with *Cep192-*biallelic variants arrested at E7 because of cell apoptosis mediated by MVA/tetraploidy cells’ acumination. Mice with heterozygous variants replicated the predisposition to male infertility. Mouse primary embryonic fibroblasts with *Cep192-*biallelic variants cultured in vitro showed abnormal morphology, mitotic arresting, and disruption of spindle-formation. In patient epithelial cells with biallelic variants cultured in vitro, the number of cells arrested during the prophase increased because of the failure of spindle formation. Accordingly, we present a novel disease gene *CEP192,* which as a link for the MVA syndrome with tetraploidy and the predisposition to male infertility.

## Introduction

Correct cell division requires chromosomal duplication and separation into two daughter cells. Errors in this process may result in multipoidy, haploid, or aneuploidy, which causes human diseases. Mosaic variegated aneuploidy (MVA) syndrome is a rare disorder, in which one-quarter or more of cells in affected individuals have an abnormal number of chromosomes [ref1-2]. Individuals with MVA syndrome exhibit microcephaly, developmental delay, and other variable abnormalities such as increased risk to malignancies and Dandy–Walker malformation [ref2]. From literatures, several patients with MVA syndrome have been reported [ref 1-7]; three disease genes have been identified for MVA syndrome, including *BUB1B* (OMIM:257300) [ref1], *CEP57* (OMIM: 614114) [ref6], and *TRIP13* (OMIM: 617598) [ref7]. Tetraploidy is another error in chromosome separation, and it refers to the presence of four copies of the genome in a cell [ref8]. In human, tetraploidy has been described frequently in spontaneous abortions[ref9] and in tumorgenesis [ref8, 10]. Genetic instability such as MVA is usually preceded by tetraploidy during tumor revolution [ref 9, 10-12]. In early time, errors in chromosome segregation have also been reported in male subfertility [ref15-16]. Variants on a centrosome gene *PLK4*, which control spindle formation and chromosome separation, has been recently reported on aneuploidy and male infertility [ref 17-20]. Nevertheless, the key molecule that drives the formation of tetraploidy is unknown; not all patients with MVA syndrome can be explained by known MVA genes, and no evidence of a molecule that link the MVA, tetraploidy, and infertility together has been reported. In this study, we report on an interesting family with two siblings suffered by MVA syndrome with tetraploidy, and other two members suffered by reduced testicular size and infertility. We exemplify a novel gene *CEP192* biallelic variants lead to the MVA syndrome plus tetraploidy, and *CEP192* monoallelic variants lead to predisposition to male infertility by a series of experiments performed on human subjects and mice models with equivalent variants.

## Results

### A family with unexplained diseases

Two siblings (patient A and patient B) that experienced microcephaly, developmental delay, limb– extremity dysplasia, facial abnormalities, and reduced testis size came to our clinic for genetic counselling (Table 1, Supplementary Methods). Genetic investigation showed that two other family members (patient C and patient D) suffered from reduced testicular volume (Supplementary Table 1), and patient D was completely infertile. Semen analysis on patient D revealed severe oligozoospermia (Supplementary Table 1). Testicular biopsy revealed a comprehensive reduction of germ cells in his available seminiferous tubules without spermatozoa (Figure 1). Similar semen parameters were observed on patient C (Supplementary Table 1). GTG-banding on patient A and B revealed that 15.52%–24.14% of lymphocytes were tetraploidy cells, 26.44%–27.27% were MVA cells, and 7%–12.33% of metaphase cells showing premature sister chromatid separation (PSCS, Figure 2, Table 1, Supplementary Figure 1, Supplementary Methods). In addition, a lower but substantial proportion of MVA/tetraploidy cells was observed on patient C and patient D (Table 1, Supplementary Methods).

**Figure 1:**
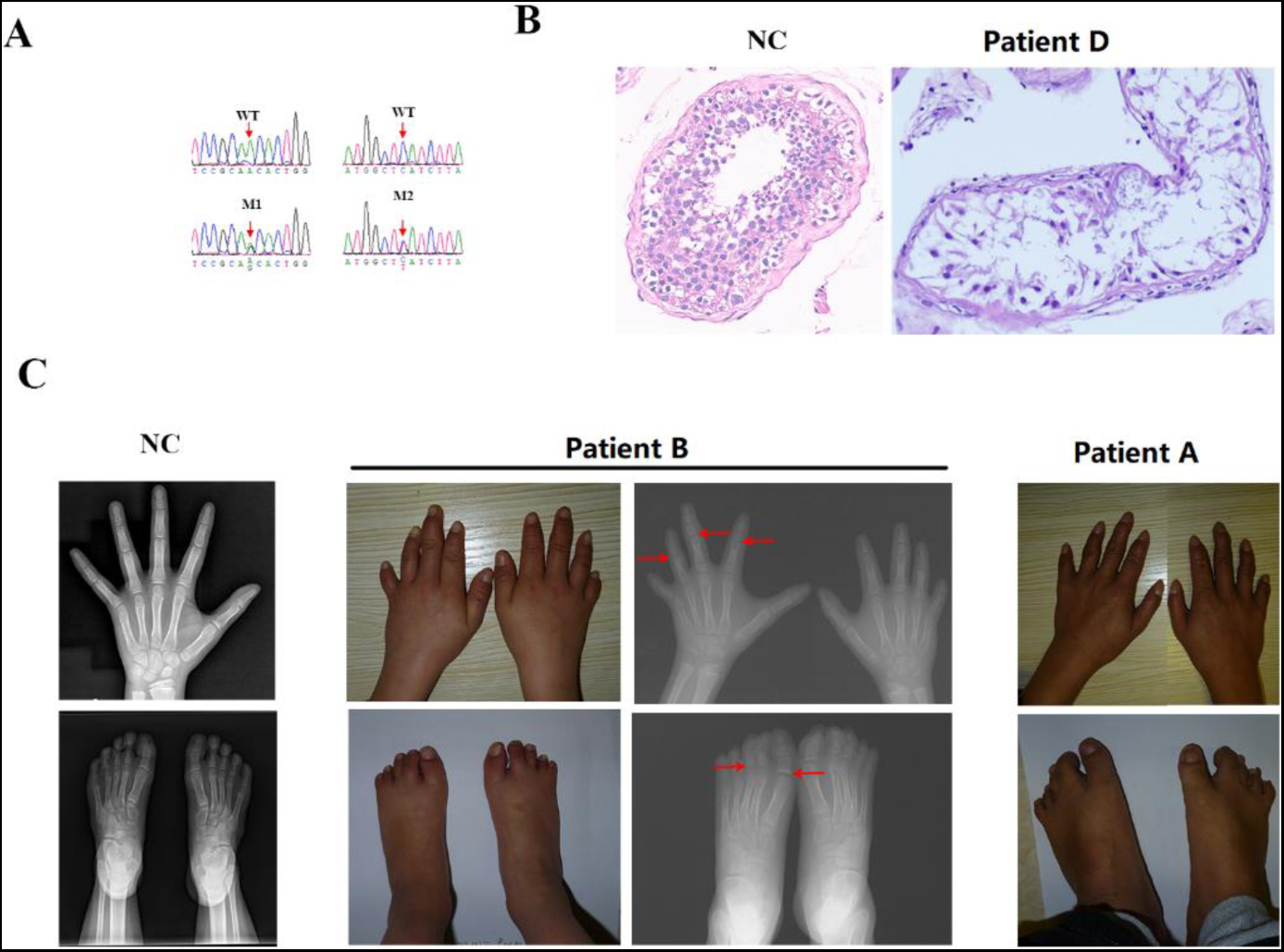
*CEP192* variants and clinical phenotypes of a family. **A:** *CEP192* variants validated by Sanger sequencing. WT mean wild type; M1 indicate variant c.5750A>G/p.N1917S; M2 refers to variant c.1912C>T/p. H638Y; P mean proband. **B:** Histological analyses of testicular tissues by H&E staining. Left: Representative normal control tubule from a 60s years-old man with prostate cancer; Right: representative tubule of patient D at 40s years old **C:** Limb extremities malformations identified by X-ray images. **NC,** healthy girl with age matched to patient B. Two siblings (patient A and patient B) have similar malformations in fingers and toes. **Fingers:** Brachydactyly; Dysplasia in middle and distal phalanges; Lack of middle phalanges of little finger; Absent epiphyseal ossification centers in the 2nd–4th middle phalanges (arrow); **Toes:** Lack of middle phalanges of the 2nd–5th toes; Syndactyly of the 2nd and 3nd toes (webbed toes); Syndactyly of the 4th and 5th toes (webbed toes); Absent epiphyseal ossification centers in the proximal phalanges of great toe and from 2nd toe to 4th toe (arrow)

**Figure 2:**
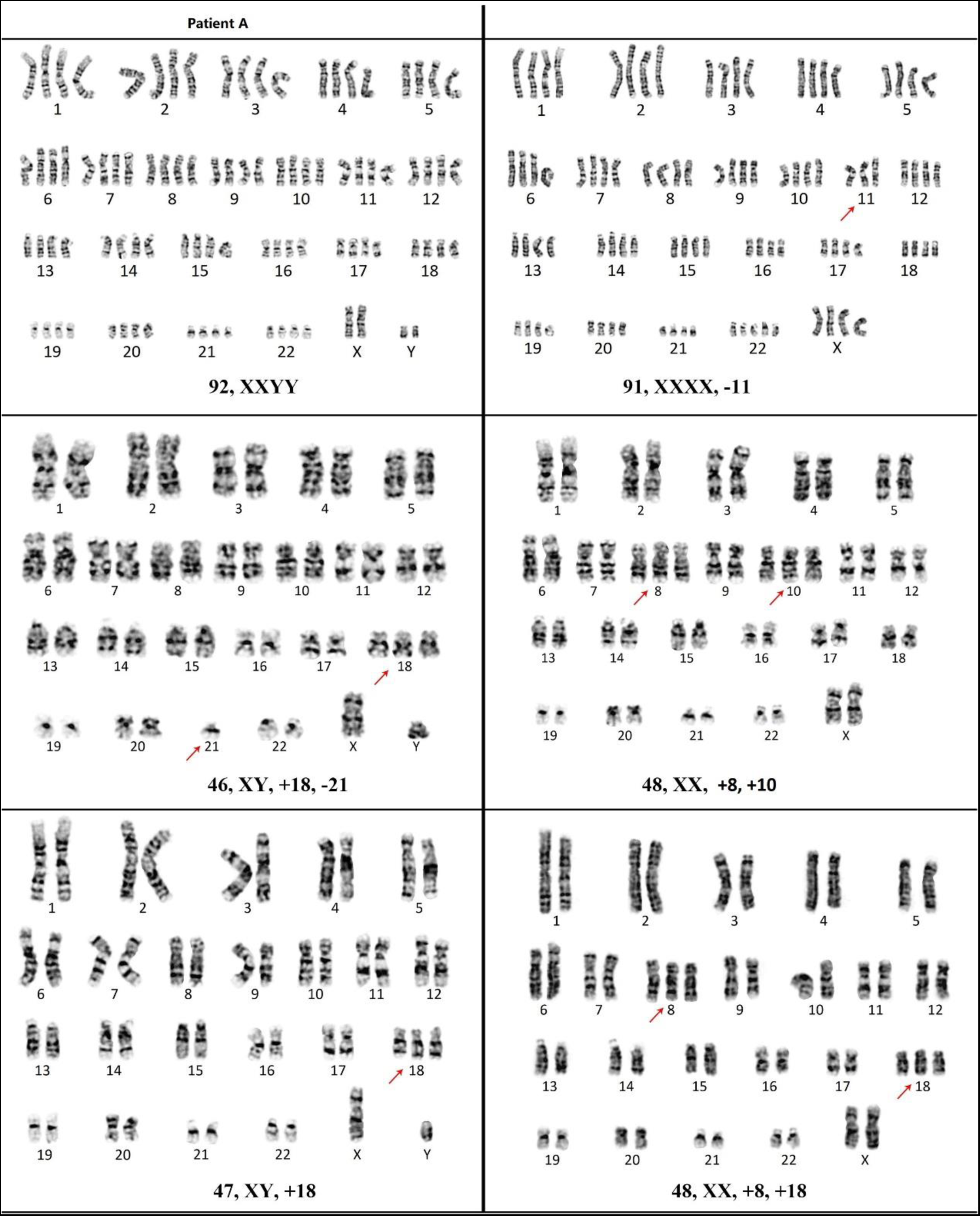
Representative metaphase cells of mosaic variegated aneuploidy and tetraploidy identified in lymphocytes of patient A and B. **Note:** Original figures are shown in Supplementary Figure 1.

**Table 1:** Cytogenetic phenotypes of the affected individuals with *CEP192* variants. F1, the index family; MVA, mosaic variegated aneuploidy. NA, not available.

| ID | *CEP192* variants | Cytogenetic phenotypes |
| --- | --- | --- |
| Patient A | Biallelic:<br>c.1912C>T/p.H638Y<br>c.5750A>G/p.N1917S | 87 cells analyzed:<br>23 cells (26%) were MVA<br>21 cells (24%) were tetraploidy |
| Patient B | Biallelic:<br>c.1912C>T/p.H638Y<br>c.5750A>G/p.N1917S | 121 cells analyzed:<br>33 cells (27%) were MVA<br>29 cells (24%) were tetraploidy |
| Patient C | Monoallelic:<br>c.5750A>G/p.N1917S | 106 cells analyzed:<br>14 cells (13%) were MVA<br>3 cells(3%)were tetraploidy |
| Patient D | Monoallelic:<br>c.1912C>T/p.H638Y | 64 cells analyzed:<br>8 cells (13%) were MVA<br>2 cells (3%) were tetraploidy |
| Patient E | Monoallelic:<br>c.5750A>G/p.N1917S | 286 cells analyzed:<br>34 cells (12%) were MVA<br>2 cell (1%) was tetraploidy |
| Patient F | Monoallelic:<br>c.5750A>G/p.N1917S | 77 cells analyzed:<br>12 cells (16%) were MVA<br>1 cell (1%) was tetraploidy |
| Patient G | Monoallelic:<br>c.5750A>G/p.N1917S | NA |

According to diagnostic criteria (a quarter or more cells were MVA) [ref1-2], and patient A and patient B fulfilled the diagnosis of MVA syndrome. Considering that, in addition to MVA, patient A and patient B also had tetraploidy, and tetraploidy has never been described in patients with MVA syndrome. Accordingly, we proposed a subtype of MVA syndrome, namely, the MVA syndrome with tetraploidy, for the siblings. To search for the cause for the family, we performed variant screening for known MVA syndrome genes (e.g., *BUB1B*, *CEP57*, and *TRIP13*) [ref 1, 6-7], but no causative variant was identified.

### Identification of *CEP192* variants

Exome sequencing (ES) was successfully performed for patient A, patient B and their parents (Supplementary Table 2). Considering that MVA syndrome is a rare, autosomal recessive disorder [ref 1, 6-7], we focused on genes with rare bi-allelic variants on patient A and patient B according to reasonable filtering strategy (Supplementary Figure 2). Only two compound heterozygous variants of *CEP192* (NM_032142.4; c.1912C>T/p. H638Y and c.5750A>G/p.N1917S) met the filtering criteria. Sanger sequencing identified the *CEP192* biallelic variants were co-segregated with syndromic phenotypes on patient A and patient B, and the *CEP192* monoallelic variant was co-segregated with reduced testicular-size/infertility on patient C and patient D (Figure 1A).

### *CEP192* associated with male infertility in general population

We determined whether *CEP192* variants were involved in male infertility in general population. Variant screening of the *CEP192* coding regions was performed for 1,264 unrelated males with idiopathic infertility. By focusing on rare variants (MaF<0.001 in Eas_gnomAD), dozens of variants were retained. The p.N1917S variant of *CEP192* was recurrently detected in three unrelated infertile males with reduced testicular volumes (patient E, F and G; Table 1). In addition, other variants on *CEP192* were repeatedly detected in infertile males (Supplementary Table 3–5). Clinical reevaluation confirmed that all three males with p.N1917S variant were infertile, and GTG-banding results revealed that a substantial part of the cells was MVA/tetraploidy (Table 1, Supplementary Method).

### Evolutionary conservation of p.N1917, and abnormal splicing of c.1912C>T

CEP192 is a 2,537 residue-long protein with eight tandem domains in its C-terminal part that are similar to members of the PapD-like superfamily [ref 21]. Tandem domains 4 and 5 of human CEP192 constitute the Spd2 domain, which is ubiquitously present in all SPD2/CEP192 homologs and represents the most conserved region among the protein [ref 21-22]. The p.N1917S variant is located in the tandem domain 5 (Figure 3A). However, the other variant, namely, the c.1912C>T/p. H638Y, is located on the N-terminal, which is not conserved among different species (Figure 3A). The “Y” amino acid can be seen in residue 638 in *Nannospalax galili* (Figure 3A). However, programs in silico prediction [ref 23-24] estimated the c.1912C>T variant affects normal splicing by disrupting Exon 14-donor splice site. Total RNA was then extracted from patient cells, and a complementary strand of DNA (cDNA) was generated. qPCR assay showed that *CEP192* cDNA was considerably reduced in cases with c.1912C>T compared with that without the c.1912C>T variant (Figure 3B). The *CEP192* transcript at the mRNA level was assessed by next-generation sequencing on a member with heterozygous c.1912C>T variant. Results showed that the c.1912T transcripts were detected in approximately 1% of the overall reads (Figure 3C). Furthermore, mini-gene assay showed that c.1912C>T led to the skipping of exon 14 (Figure 3D), resulting in a frame shift and a premature termination codon (p.P604E*2). Thus, the functional mechanism of c.1912C>T should be the vast majority of mutant transcripts undergoing abnormal splicing, and the shortened transcripts mediated nonsense-mediated mRNA decay.

**Figure 3:**
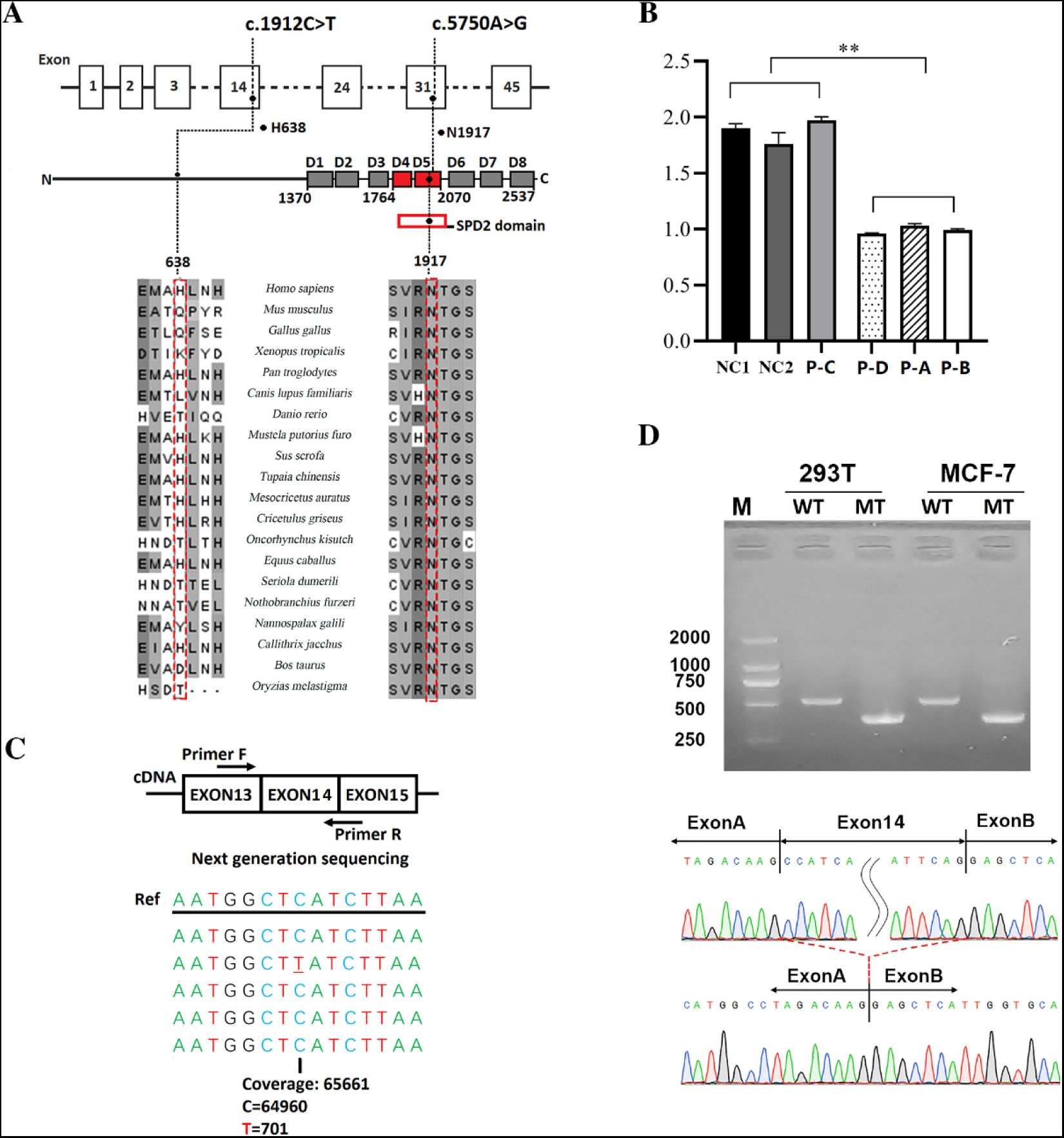
Functional evaluation of *CEP192* variants. **A:** Location of two variants (c.1912C>T/p. H638Y and c.5750A>G/p.N1917S) in *CEP192* gene (upper) and in the known domains of protein (middle). Sequence alignment of CEP192 residue 638 and residue 1917 across different species (bottom). **B:** Expression level of *CEP192* mRNA in individuals with or without c.1912C>T variant. The relative abundance of *CEP192* mRNA expression was calculated by normalization to ACTB level. **Group 1:** Individuals without *CEP192* c.1912C>T (NC1 and NC2 were two unrelated healthy individuals; Patient C (P-C) was a family member with c.5750A>G but without c.1912C>T). **Group 2:** Individuals with *CEP192* c.1912C>T variant (Patient A, B and D). The statistical significance of the departure of the observed ratio from the expected ratio is represented by \*\**p* < 0.01. **C:** Composition analysis of c.1912C>T transcripts in lymphocyte cells of patient D by next-generation sequencing **D:** Minigene assay for c.1912C>T variant of *CEP192*. **Upper:** PCR products amplified from RT-PCR products (from HEK293T and MCF-7 cells, respectively) were separated by electrophoresis. Approximately 500 bp fragment was identified in the pcMINI-CEP192-WT cells, and a small fragment was identified in pcMINI-CEP192-MT cells (in both HEK293T or MCF-7 cells); **Bottom:** Sanger sequencing of the RT-PCR products illustrated the skipping of Exon 14 (whole exon 14, 127 bp) in mutant status. WT means wild type; MT means *CEP192-*c.1912C>T variant; M means ladder.

### Animal models confirmed that both *CEP192* variants are pathogenic

Two lines of *Cep192*-edited mice were generated consisting of a knock-in mice (*Cep192*^+/M^) with a variant equivalent to human N1917S and a knock-out mice (*Cep192*^+/-^) mimicking the haploinsufficiency effect of the human c.1912C>T splicing variant (Supplementary Figures 3–4). In progeny derived from the crosses between two *Cep192*^+/-^ mice, the ratio of *Cep192*^+/+^ to heterozygote (*Cep192*^+/-^) was close to 1:2 (34 litters, 131 offspring calculated, *Cep192*^+/+^=52, *Cep192*^+/-^ =79). No *Cep192*^-/-^ offspring was obtained (Figure 4A, Supplementary Table 6), suggesting embryonic lethality in the *Cep192*^-/-^ state. In parallel, in the progeny derived from the crosses between two *Cep192*^+/M^ mice, the ratio of *Cep192*^+/+^ progeny to *Cep192*^+/M^ was close to 1:2 (21 litters, 158 offspring calculated, *Cep192*^+/+^=64, *Cep192*^+/M^ =94). No *Cep192*^M/M^ offspring was obtained (Figure 4A, Supplementary Table 7). Subsequently, *Cep192*^+/M^ mice were bred with *Cep192*^+/-^ mice to further verify the causality of both variants. The *Cep192*^+/+^*, Cep192*^+/-^, and *Cep192*^+/M^ progeny can be survivable. However, no *Cep192*^-/M^ mice were obtained (Figure 4A, Supplementary Table 8). The results above implicated that both the knock-out and the N1917S allele were causative agents.

**Figure 4:**
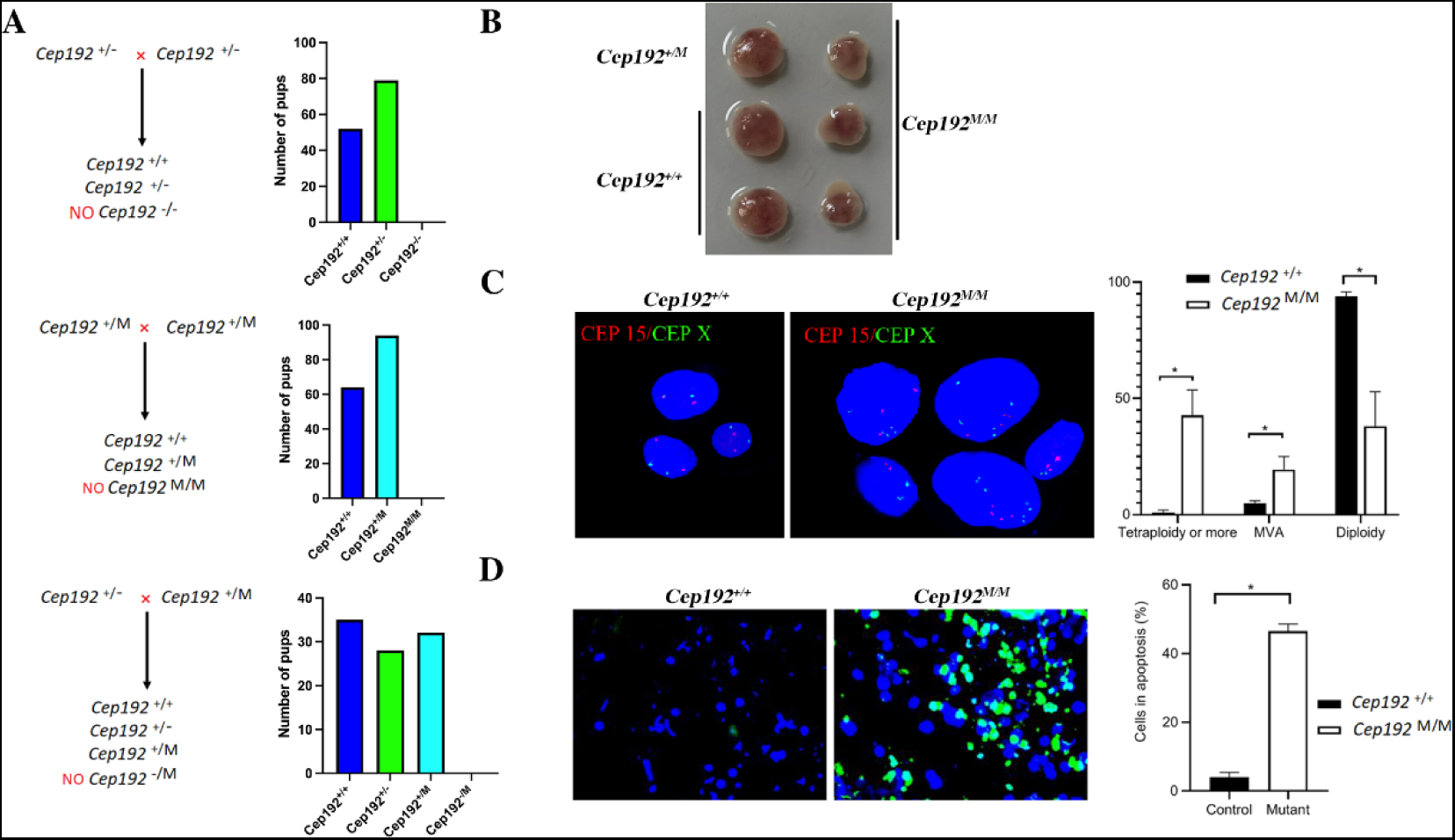
Genetic analysis performed on mouse models. **A:** Genotype analysis of progeny from the heterozygous intercrosses. **B:** Representative embryos with different genotypes in E10. *Cep192*^M/M^ embryos arrested at about E7. **C:** Representative interphase nuclei monitored by fluorescence in situ hybridization. **Left:** Cells from wild-type embryos, three cell nuclei stained by two red (chromosome 15) and two green (chromosome X) signals; **Middle:** cells from mutant embryos (upper left, three red, and five green signals; upper right: five green and four red signals; lower left: two green and two red signals; lower middle: four green and four red signals; lower right: three red and two green signals); **Right:** Count of normal diploidy cells, MVA, and tetraploidy cells for *Cep192*-mutant or wild-type embryo cells. For each sample, randomly 100 cells were analyzed (*n*=3). **D:** Apoptosis (cells stained by green) in cells from wild-type or *Cep192^M/M^* stagnated embryos (n=3)

### Mutant mice replicated MVA and tetraploidy phenotypes

In vitro fertilization was performed. *Cep192* homozygous mutant embryos could develop into blastocysts in vitro (Supplementary Figure 5). To explore the stage(s) in which the homozygous embryos undergo arrestation, we sacrificed pregnant mice at various days post coitum. Embryos were dissected at various developmental stages. *Cep192*^-/-^ embryos appeared morphologically normal at E7 compared with their *Cep192*^+/-^ or *Cep192*^+/+^ littermates (Supplementary Figure 6). Embryonic death was detected for *Cep192*^M/M^ embryos at E10, the size of dead embryos is equivalent to that in E7 (Figure 4B). The same results were observed on *Cep192*^-/-^ embryos (Supplementary Figure 7). Embryos were dissected and ground to cell suspensions. Fluorescence in situ hybridization (FISH) showed a significant increase of MVA and tetraploidy cells in *Cep192*^M/M^ embryo cells (Figure 4C). TUNNEL assay revealed that the number of apoptotic cells increased in *Cep192* ^M/M^ embryos compared with that of *Cep192*^+/+^ embryos (Figure 4D). Similar results were obtained in cell samples from *Cep192*^-/-^ embryos (data not shown). In addition, when *Cep192^+/-^*or *Cep192^+/M^* males were mated with wild type (WT) females, approximately 10% of the mated females experienced dystocia. The uteri of females with dystocia were dissected. Fetuses arrested in about E13-14 were observed; all arresting fetuses were heterozygous (23 tested, 10 were *Cep192*^+/-^; 13 were *Cep192*^+/M^) (Supplementary Figure 8). Similar to *Cep192*^-/-^ or *Cep192*^M/M^ dead embryos, a remarkable proportion of cells (from the *Cep192*^+/-^ or *Cep192*^+/M^ arrested embryos) were MVA or tetraploidy (Supplementary Figure 9). TUNNEL assay also revealed that up to 40%–50% of the cells from the *Cep192*^+/M^ that arrested embryos underwent apoptosis (Supplementary Figure 9).

### Male mice with *Cep192* heterozygous variants replicated infertility

Breeding assays were performed to check the reproductive capability of male mice with Cep192 heterozygous variants. *Cep192*^+/-^ males were mated with WT females from complete sexual maturity (2 months) to 9 months. In comparison with WT males mated with WT females with average number of litters of 6.6, the *Cep192^+/-^* or *Cep192 ^M/-^* males mated to *WT* females exhibited reproductive defects with phenotypic variation among individuals (Figure 5; Supplementary Table 9-11). The number of litters or pups was reduced considerably in *Cep192*^+/-^ or *Cep192 ^+/M^* males when they mated with *WT* females (Figure 5AB; Supplementary Tables 9–11). Histological analysis was performed. In the majority of the fertile heterozygotes, testicular size and histology appeared normal. However, the testes of the infertile *Cep192^+/-^* or *Cep192^+/M^* males were smaller than those of the *Cep192*^+/+^ littermates (Figures 5CD, Supplementary Table 12). H&E staining revealed that in comparison with a large number of sperms shown in the epididymides of *Cep192*^+/+^males, the epididymides of the infertile heterozygous mice were nearly empty (Figure 5 E*). Meanwhile, in comparison with Cep192*^+/+^ littermates*, a substantiate reduction of* germ cells was observed in the seminiferous tubules of infertile *Cep192*^+/-^ or *Cep192 ^+/M^* males (Figure 5 E).

**Figure 5.**
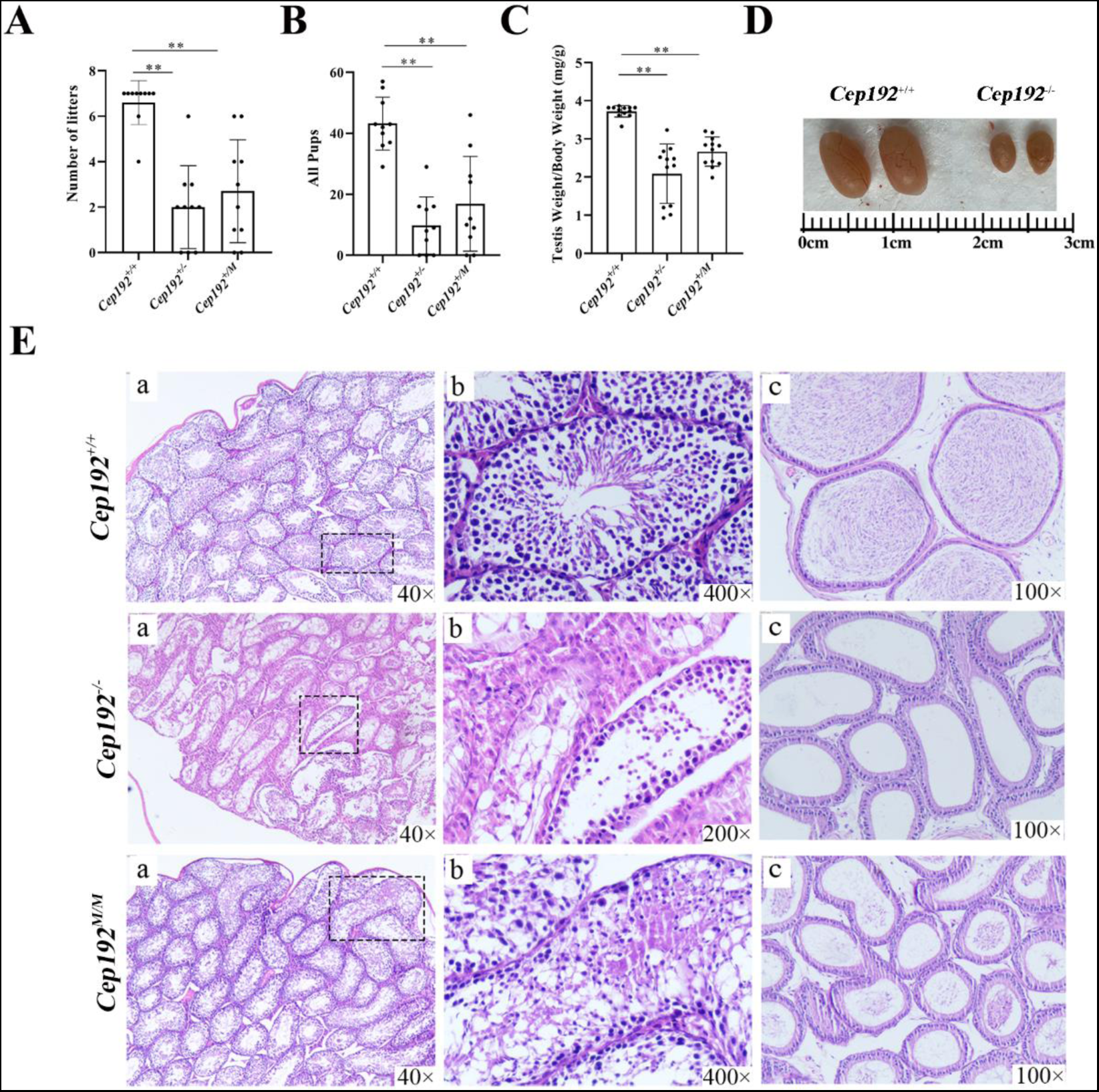
Reproductive phenotypes of infertile male mice with *Cep192* heterozygous variant. **A–B:** Number of litters and total pubs from the persistent mating of *Cep192^+/-^*, *Cep192*^+/M^, or wild-type males with wild-type females from complete sexual maturity (2 months) to 9 months. **C:** Testes to body weight ratios of infertile heterozygous mice (*Cep192*^+/-^, *Cep192*^+/M^) and wild-type mice at 180 days. **D:** Representative figure of testes between a *Cep192^+/-^* mice with infertility and a wild-type mice at 180 days. ***E:*** H&E staining of testes (a and b) and epididymides cauda (c) from wild-type mice and infertile heterozygous mice (*Cep192*^+/-^, *Cep192*^+/M^) at 180 days old.

### Spindle formation was disrupted in *CEP192-*mutant cells

Mice embryo cells were cultured in vitro. Only a few *Cep192*^-/-^ or *Cep192*^M/M^ cells can adhere to disk walls (less than 1% vs. *Cep192*^+/+^ 60%). For the cells that can adhere, the cell division was very slow (almost cannot divide). As shown in cells in the interphase, the mutant cells exhibited bipolarized (or dysmorphic) cell shape (Figure 6). In comparison with *Cep192*^+/+^ cells, an appropriate number of microtubules (MTs) were arranged in an orderly manner around the centrosome, the *Cep192*^-/-^ or Cep192 ^M/M^ cells exhibited decreased volume of MTs, and the MTs were disorganized (Figure 6AB). Some of the mutant cells seems lacking microtubule organizing center (MTOC, Figure 6AB). In several cells likely in mitosis, chromosomes arranged around lacking a spindle formation (Figure 6B). The investigation was focused on human cells with *CEP192* variants to check the spindle status in mitotic cells. Epithelial cells (from patient B) were cultured in vitro. In the prophase, the nucleation of MTs occurred randomly, leading to disorganized, non-bipolar structures of spindle (Figure 7). Quantitative analysis was performed and indicated that the *CEP192*-mutated cells exhibited an increased proportion of mitotic cells in prophase (81.90% vs. 20.38% in control cells, Figure 7EF). We propose here that cells lacking normal spindle structure have high probability to be developed to the MVA or tetraploidy cells.

**Figure 6:**
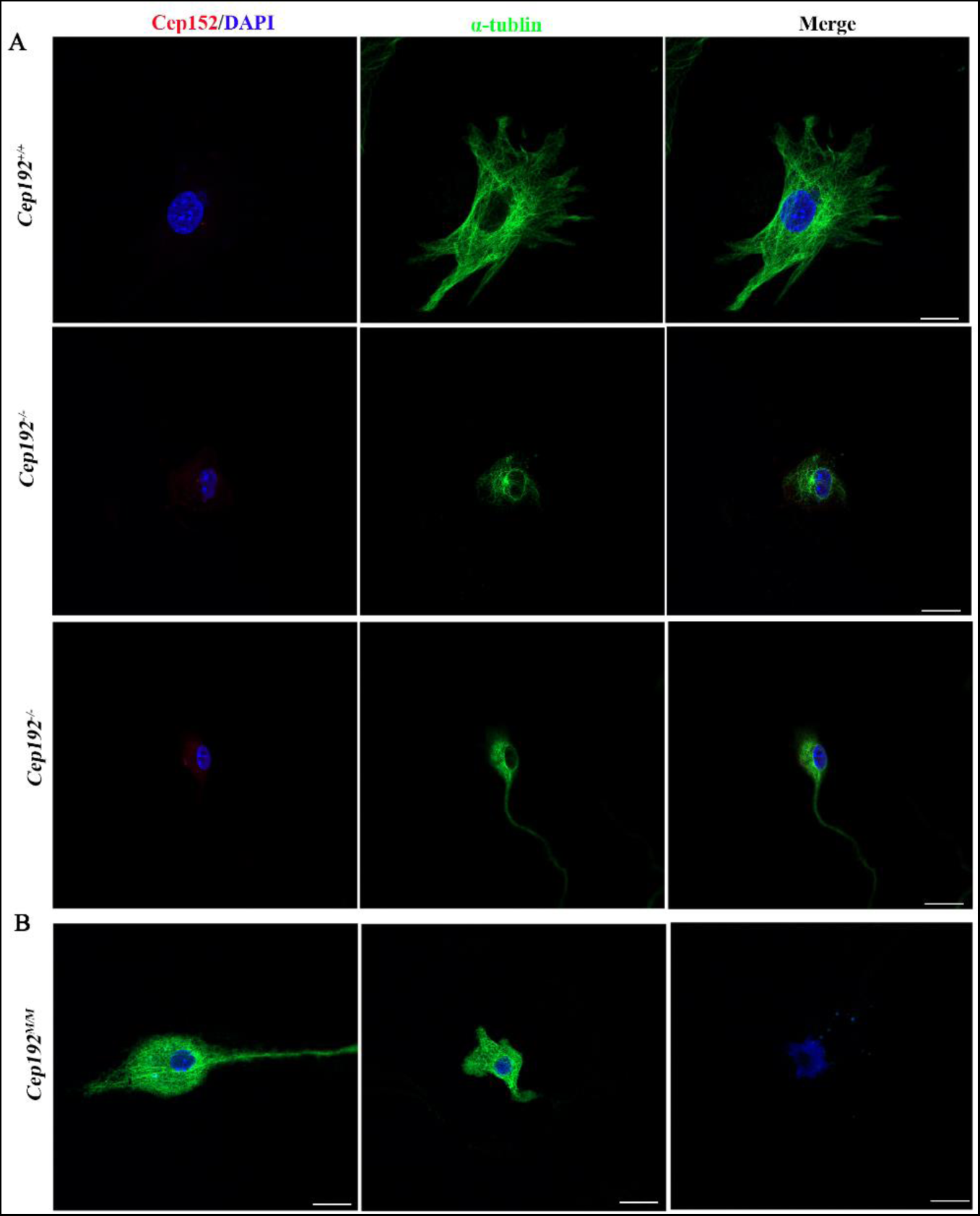
Alteration in mouse primary embryonic fibroblasts without functional CEP192. A: Immunofluorescence staining of primary embryonic fibroblasts from wild-type (Cep192^+/+^) and knock-out (Cep192^-/-^) mice by using antibodies against a centrosome marker protein CEP152 (red), microtubules (green), and DAPI staining of DNA (blue). Top: CEP152 was co-localized with the microtubule organazing center (MTOC) in Cep192^+/+^ cell; Middle: one CEP152 signal was co-localized with MTOC in Cep192^-/-^cell, but the other mutant was not; Bottom: two CEP152 signals were located in cytoplasm that distal to the nuclear (obviously not in MTOC) and a CEP152 signal in MTOC in a uni-polarized cell. Scale bars, 7.5 µm. B: Immunofluorescence staining of primary embryonic fibroblasts from knock-in (Cep192 ^M/M^) mice by using antibodies against microtubules (green) and DAPI staining of DNA (blue). Left: a bipolar cell; Center: an abnormal multipolar cell; Right: cell in mitosis but without spindle and without microtubule. Scale bars, 7.5 µm.

**Figure 7:**
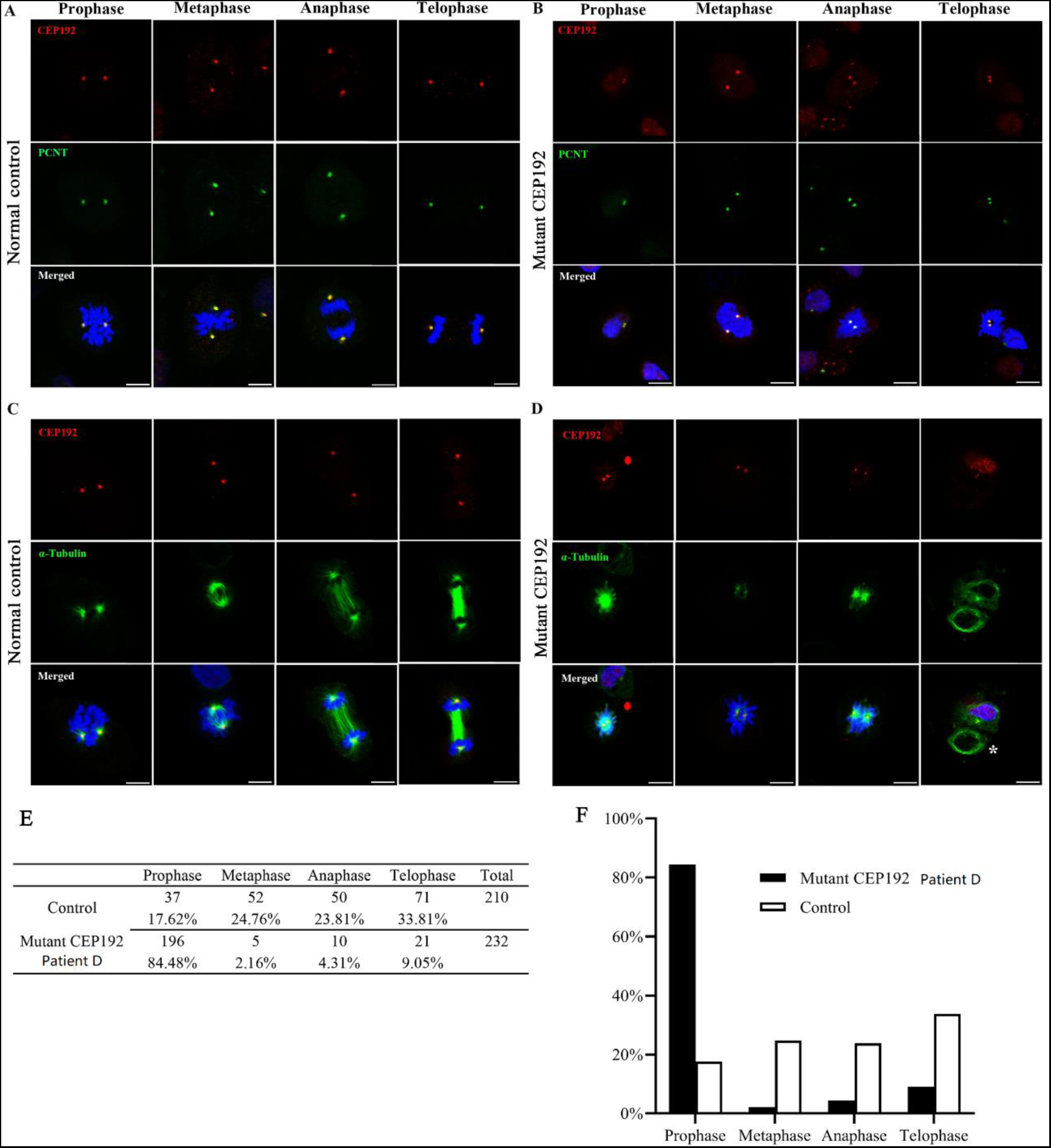
Mitosis defects and spindle formation abnormalities in cells from a patient with *CEP192* biallelic variants. **A–B:** Immunofluorescence staining of epithelial cells from a patient patient B and normal control using antibodies against CEP192 (red) and PCNT (green), and DAPI staining of DNA (blue). CEP192 proteins were co-localized with the centrosome in the different stages of mitosis in both cells from normal control and patient B. Scale bars, 7.5 µm. **C–D:** Immunofluorescence staining of epithelial cells from patient (patient B) and a normal control using antibodies against CEP192 (red) and α-tubulin (green), and DAPI staining of DNA (blue). **C:** Normal spindle formation at interphase, metaphase, anaphase, and telophase in epithelial cells were observed in the normal control. **D:** Two centrosomes were not fully pulled apart per nucleus in epithelial cells from patient. **Asterisk** indicates the abnormal cells with unevenly distributed nuclei during cell division. Scale bars, 7.5 µm. **E–F:** Quantification of different stages in mitosis in epithelial cells from normal control and patient B.

## DISUSSION

In present study, we have investigated a family (F1) with two siblings that suffered from developmental delay, microcephaly, limb extremities malformation, and reduced testicular size. In addition, two other male family members showed reduced testicular size/infertility. Genetic inheritance was initially unknown for the family. The performance of GTG-banding identified MVA and tetraploidy cells in the patients. ES and further validation disclosed that *CEP192* biallelic variants associated with the MVA syndrome with tetraploidy, and the *CEP192* mono-allelic variants associated with reduced testicular size/infertility. From 1,264 unrelated patients, the same heterozygous *CEP192* variant (in family F1) and other variants were associated with male infertility, confirming that *CEP192* is involved in male infertility. Accordingly, two mice models with equivalent variants were generated. Mutant mice replicated MVA/tetraploidy cytogenetic phenotypes and the susceptibility to male infertility. In Online Mendelian Inheritance in Man (616426), *CEP192* is a protein coding gene without a clear link to human disease. Thus, the present study should represent the identification of a novel disease-gene *CEP192* that links to two human diseases, the MVA syndrome with tetraploidy and male infertility.

*CEP192* encodes a centrosome protein. Centrosomes are organelles that serve as the MTOC for animal cells [ref25-27]. Experiments in vitro by RNA interference indicated that in the M phase, CEP192 is important for spindle formation [ref25]; in the interphase, CEP192 plays an important role in MT organization (when it depleted, cells became “slim shape”)[ref28]. In the present study, by focusing on MVA/tetraploidy cellular phenotypes, we identified *CEP192* natural variants associated with human disorders. Based on the cells from both human and mice, cells in M phase with *CEP192* biallelic variants exhibited disorganized, non-bipolar structures of the spindles; in the interphase, mice fibroblasts with bi-allelic variants exhibited an elongated, bipolar, unipolar, or multipolar cellular shape. Therefore, the present study proved that CEP192 was vital for spindle formation (in mitosis) and normal MT organization (in interphase) from a new perspective.

Notably, the different embryonic development fate was observed between human and mice with *CEP192* biallelic variants. Mouse embryos with biallelic *Cep192* variants (*Cep192^M/M^ Cep192^M/-^ Cep192^-/-^*) were arrested at about E7, whereas human with *CEP192* compound heterozygous variants (c.1912C>T/p. H638Y and c.5750A>G/p.N1917S, i.e., the patient A and patient B) were survivable. Such situation may result in subtle differences in one of variants between humans and mice. *Cep192^M/M^* mice variant (c.5675 A>G/N1892S) was equivalent to human c.5750A>G/p.N1917S variant, while the *Cep192^-/-^* mice genotype (complete knock out) was different to human c.1912C>T/p. H638Y. In mouse model generation, considering that c.1912C/p. H638 (of human) is not conserved among different species, but majority portion of c.1912T transcripts was undetectable in patient-RNA sequencing (as abnormal spicing), the *cep192*-knockout mice was generated to mimic the effects of the c.1912C>T/p. H638Y variant. However, unlike the complete knock out in mice, approximately 1% of c.1912T transcripts can be detected in patient cells (Figure 3C). Therefore, approximately 1% of the c.1912T transcripts (that escaping exon 14 skipping) conserve a few CEP192 function that drives cell mitosis. Based on a previous study, U2OS cells with 5% CEP192 function can reserve the capacity to divide, but the cell division and migration were partially blocked [ref 28]. Thus, we proposed here that the complete depletion of *CEP192* leads to the failure of cell division, indicating that CEP192 is indispensable in cell mitosis, and cells that conserve a few CEP192 function can divide, but the division speed is slowed down.

Faithful segregation of homologous chromosome and sister chromatid is essential for generating functional spermatozoa [ref 29]. Chromosome segregation errors during meiosis may result in spermatocytes apoptosis, aneuploidy, or tetraploidy and finally cause azoospermia or oligozoospermia and male infertility [ref 30]. CEP192 works as a distinct scaffold to recruit PIK4 to centrosomes [ref 27]. The loss of the CEP192-dependent interaction with PLK4 resulted in impaired centriole duplication, thus delaying cell proliferation [ref 27]. *PLK4* is a known gene for a recessive disorder with partial MVA-related phenotypes [ref 17]. In heterozygous status, *PLK4* variants cause male hypogonadism, azoospermia, and germ cell loss in humans and mice [ref18-20]. In the present study, both human and mice with *CEP192* heterozygous variants showed susceptible to infertility is probably the mutant CEP192 fails to recruit PLK4.

This has some limitations. In present study, approximately 2030% of male mice with monoallelic *Cep192* defect (*Cep192*^+/-^ or *Cep192*^+/M^) were identified to be infertile, from which small testes and histologic changes were observed. However, the remaining fertile *Cep192*^+/-^ or *Cep192*^+/M^ male mice exhibited seemingly normal testicular histology. As the embryonic lethality of the *Cep192*^-/-^ or *Cep192*^M/M^ mice, the effects in testicular change could not be studied in the homozygous state. Loss of germ cell and reduction in testis size present in a part of the *Cep192*- heterozygotes mice may be a partial phenotype. The effect of a homozygous *Cep192* in the testis could be investigated using a conditional knockout model in our next work.

In conclusion, the present study has identified a novel disease gene *CEP192*, which links a rare (the MVA syndrome with tetraploidy) and a common disorder (male infertility). These findings expand our knowledge of the relationship between centrosome proteins and human disease and will allow the precise genetic diagnosis of MVA syndrome and male infertility.

## Supporting information

supplementary methods, tabs and figs

## Data Availability

All data produced in the present study are available upon reasonable request to the authors

## AKNOWLEDGMENTS

We thank all family members who participated in this study. National Natural Science Foundation of China (31501017 to Yongjia Yang; 82101961 and 82171608 to Yue-Qiu Tan, 82201773 to Wen-bin He; 81701437 to Fen Tian), Natural Science Foundation of Hunan Province (2021JJ30390 to Yongjia Yang; 2023JJ30716 to Wen-bin He), the National Key Research and Development Program of China (2022YFC2702604 to Yue-Qiu Tan) and the Hunan Health Commission Research Fund (B2019019 to Yongjia Yang; B202301039323 to Wen-bin He). We would like to thank the technician Yongzhong Guo for his contribution in animal feeding and maintenance. We would like to thank the technician Yongzhong Guo for his contribution in animal feeding and maintenance.

## AUTHOR CONTRIBUTIONS

The project was conceived and the experiments were planned by Yongjia Yang. Experiments about sequencing of *CEP192* for 1264 infertility males was planned by Yue-qiou Tan. The review of phenotypes and the sample collection for the index family were performed by Yongjia Yang, Jihong Guo, Ming Tu, Liu Zhao, Xinghan Wu and Hua Wang. The review of phenotypes and the sample collection for the 1264 males with infertility were performed by Wen-bin He, Yue-Qiu Tan, Chen Tan, Lan-Lan Meng, Guang-Xiu Lu, Ge Lin, Yue-Qiu Tan. Comprehensive cytogenetic analysis was performed by Yongjia Yang, Jihong Guo, Fang Shen, Ming Tu, Liu Zhao, Chen Tan. Gene functional experiments were performed by Lei Dai, Yongjia Yang, Wen-bin He, Fen Tian, Fang Shen, Mei Deng, Shuju Zhang. Bioinformatic analysis was performed by Yu Zheng, Lan-Lan Meng, Yu Peng and Yongjia Yang. Animal experiments were performed by Jihong Guo, Wen-bin He, Zhenqing Luo, Fen Tian, Wei Liu and Yongjia Yang.

## EXPERIMENTAL PROCEDURES

### Human subjects

Several members in a family were recruited to present study. A total of 1,264 idiopathic infertile men with azoospermia, oligozoospermia, and severe oligo-asthenospermia were recruited. ES data of such cohort of patients with idiopathic infertility have been described previously [ref 31, 32]. The present study was approved by the ethics committee of the Hunan Children’s Hospital (for the index family, approval number: HCHLL201558, Changsha City, Hunan Province, China) or by Institute of Reproductive and Stem Cell Engineering, Central South University (for 1,264 males with infertility, approval number: LL-SC-2017-025 or LL-SC-2019-034, Changsha City, Hunan Province, China). Appropriate written informed consent was obtained from participated subjects or the guardians of the minors.

### G-bands by trypsin using Giemsa (GTG banding)

The peripheral venous blood of the patients and their family members was collected in a vacutainer sodium heparin vial. Slides were prepared from phytohemagglutinin-stimulated peripheral lymphocyte cultures according to standard cytogenetic methods. In brief, 0.4 mL of whole blood was cultured in lymphocyte medium (5 mL) for 68 h. Then, colchicine (50 µl, 20 µg/mL) was added 2 h before cell harvesting. GTG banding at a 400–500 band level was performed in accordance with the standard laboratory protocol. Two cultures corresponding to two series of slides from each sample were separately prepared and analyzed. At least 40 metaphases were analyzed for each individual. To confirm that the MVA/tetraploidy/PSCS was not occasional, we performed GTG banding for several individuals with interests.

### Histological analysis

Testes tissues were fixed in Bouin’s solution overnight and then embedded into the paraffin. After the tissue-blocks were sectioned (5 mm thickness), the tissue slides were generated and deparaffinized by xylene. Then the slides were rehydrated by gradient ethanol, sequentially stained with hematoxylin/eosin, and sealed with neutral resin. The images were captured under a microscope (Olympus BX51, Tokyo, Japan).

### ES and variant validation

Genomic DNA from peripheral blood samples was extracted using a QIAamp DNA blood midi kit (Qiagen, Hilden, Germany) according to the manufacturer’s protocol. ES was performed on four family members as described previously [ref33-34]. Specific PCR primers flanking the suspected variants of *CEP192* were used to amplify the region, including Exon14F: 5’- ttgatgtcatttagctgttttacca-3’, Exon14R: 5’-ccatttaacccctaagttacgc-3’, Exon31F: 5’-gcaatcttattttggaaggcgt-3’, and Exon31R: 5’-cgacacagacacaagtgcat-3’. The purified PCR products were sequenced on a genetic analyzer (Applied Biosystems, A3500, CA, USA).

### Total RNA preparation and real-time PCR assay

Peripheral blood mononuclear cells (PBMCs) were separated from peripheral venous blood, and then cultured in Gibco RPMI 1640 medium (11530586, Thermo Fisher Scientific, USA) with phytohemagglutinin-stimulation for 48 h. Total RNA was extracted from approximately 1 × 10^6^ PBMCs for each individual by using TRIzol™ (15596026, Thermo Fisher Scientific, USA) according to the manufacturer’s instructions. First-strand cDNA was synthesized using RevertAid first strand cDNA synthesis kit (K1622, Thermo Fisher Scientific, USA). Real-time PCR was performed using SYBR® Green Premix Pro Taq HS qPCR kit (AG171101, Accurate Biotechnology, China) on Roche LightCycler 480 II (Switzerland). The following primers (Forward: 5’- GTTGCCTTGGTGGTGGTAAC-3’; reverse: 5’-GTGCCTGGGACTGTTCATTT-3’, predicted size: 186 bp) were used for *CEP192*, and primers (5’-ATGGGGAAGGTGAAGGTCG-3’, Reverse 5’- GGGGTCATTGATGGCAACAATA-3’, predicted size: 108 bp) were used for GAPDH (as an internal control). All primers were synthesized by a local biology company (BGI, China).

### Minigene analysis

Minigene analysis was performed to investigate whether the c.1912C>T variant of *CEP192* (exon 14) affect the pre-mRNA splicing in vitro [ref 35]. Amplified-target-fragments generated by standard overlapping PCR using the genomic DNA of the patient (II:2) and digestion were cloned into the plasmids vector. The recombinant vectors (WT and MT) were then transiently transfected into MCF-7 and HEK293T cells by using low-toxicity Lipofectamine following the manufacturer’s instructions (Life Technologies, Carlsbad, CA, United States). Cells were harvested 48 h after transfection, and total RNA was extracted with TRIzol (15596026, Thermo Fisher Scientific, USA). Then, cDNA is synthesized by reverse transcriptase by using a reverse transcription kit (K1622, Thermo Fisher Scientific, USA) according to the manufacturer’s instructions. RT-PCR was performed using specific primers to amplify plasmids containing the exon 14 of *CEP192.* The primers used in the experiments are shown in Supplementary Table 13. The construction of recombinant plasmids is shown in Supplementary Figure 10.

### Amplicon sequencing

The human c.1912C>T/p. H638Y variant was located on exon 14 of *CEP192*. A pair of primers that cover exon 13 and exon 15 was designed (forward: 5’-GTTGCCTTGGTGGTGGTAAC-3’; reverse: 5’- GTGCCTGGGACTGTTCATTT-3’, predicted size: 186bp). The fragment was amplified using patient’s cDNA with the predicted size of 186 bp. The PCR product was subsequently sequenced on an Illumina platform (Illumina Inc., San Diego, CA, USA) to detect the frequency of c.1912T transcript.

### Generation of *Cep192* mutant mice

Two mice models were generated on C57BL/6Nby CRISPR/Cas9-mediated genome engineering (Cyagen Biosciences, Suzhou, China), including a knockout (*Cep192* ^+/-^) and a knock in (*Cep192* ^+/M^) mice. Exons 6–41 of *Cep192* were deleted from the knock out (*Cep192* ^+/-^) mouse by using the single-guide RNAs (sgRNA-1: GGCGCTGGGCCTCTTTAAAGTGG, sgRNA-2: GGAGACAAACTCAAGTGACGAGG), which is equivalent to that observed in patients (c.1912C>T). For generating knock in (*Cep192* ^+/M^) mouse, a single-guide RNA (sgRNA-3: GTTTTTTCTATTCGCAACACTGG) was designed to target c. 5675 A>G/N1892S, which was analogous to the variant p.N1917S in patients. All experiments involving animals were approved by the institutional animal ethics committee of Hunan Children’s Hospital. The strategy and primers sequences used for genotyping the engineered mice are provided in Supplementary Figure 3-4.

### *In vitro* fertilization in mice

In vitro fertilization (IVF) was conducted in *Cep192* knock-out mice, as described previously [ref 36]. In brief, female mice were superovulated via injection with 10 IU of pregnant mare serum gonadotropin, followed by injection of 10 IU of human chorionic gonadotropin (hCG, Livzon) 48 h later. After 15–16 h, sperm samples collected from mouse cauda epididymides were added into HTF drop (EasyCheck, M1150). Next, cumulus-intact oocytes collected from superovulated female mice were transferred into a sperm-containing fertilization drop. After incubation for 5 h, mouse embryos were washed in another fertilization drop and transferred into the M16 medium (M7292, Sigma-Aldrich) for further culture (37C, 5% CO_2_). The fertilization rates were evaluated by recording the number of two-cell embryos and blastocysts at 20 and 96 h later. For genotyping of blastocysts, and whole genome amplification (WGA) was performed using the REPLI-g single cell WGA kit (Cat. No.150343), and the following PCR primers and genotyping parameters are provided in Supplementary Figure 3.

### Fluorescence in situ hybridization in mice cells

The embryo-cells of the *Cep192* mice were analyzed by (FISH and compared with a control sample (wild type mice in the same litter) using a previous reported method [ref 37]. Briefly, the mice embryos were dissected and grinded to single-cell suspension, the suspension was spread on slides, and the cells were fixed, denatured, and hybridized with two specific probes for mouse chromosomes 15 and X (Future Biotech Co., Ltd.). Subsequently, the slides were rinsed, counterstained, and imaged via fluorescence microscopy.

### Immunofluorescence analysis

Cells were plated in gelatin coated 24-well chamber slides. After 48 h post-plating, the cells were washed with DPBS and fixed for 20 min with 4% paraformaldehyde. After washing, the cells were permeabilized with 0.1% Triton X-100 for 15 min and blocked for 30 min with 5% bovine serum albumin (BSA). Cells were then incubated with first antibodies in 5% BSA at 4 °C overnight. Cells were washed several times with PBST. After incubating with secondary antibodies for 1 h, the samples were cover slipped with DAPI. Slides were examined and images were captured under the confocal fluorescence microscopy.

For human cells, rabbit anti-human CEP192 (Proteintech, 18832-1-AP, 1:50) and mouse anti-human PCNT (Abcam, ab28144, 1:100) were used to analyze the centrosome formation CEP192 colocalization; rabbit anti-human CEP192 and mouse-anti human α-Tubulin (Cell Signaling Technology, #3873S,1:100) were used to analyze spindle formation. The stage frequency of spindle formation was analyzed by analyzing at least 100 nuclei per slice via randomly selected field from each slice by two independent researchers who were blinded to the status. For mouse embryo cells, rabbit anti-mouse CEP152 (GeneTex128027, 1:100) was used to analyze the centrosome location of a cell. Mouse-anti mouse α-tubulin (Cell Signaling Technology, #3873S,1:100) was used to stain microtubules.

### TUNEL assay

Terminal deoxynucleotidyl transferase dUTP nick-end labelling (TUNEL) assay (DeadEndTM Fluorometric TUNEL System, Promega, Madison, WI, USA) was performed, according to the manufacturer’s instructions, on the cells collected from the Cep192-mutant mice-embryos (both the homozygous or heterozygous genotype) and from wild type mice-embryos (from the same litter).

### Mouse embryo-cells cultured *in vitro*

E9 embryos (Cep192 ^M/M^, Cep192^-/-^ and wild-type) were used to generate mouse embryonic fibroblasts. Briefly, the whole embryo was minced and trypsinized in 0.25% trypsin for 15 min at 37 °C. Cells were released through mechanical trituration and grown in Dulbecco’s modified Eagle medium, 10% fetal calf serum, and 100 μg of penicillin–streptomycin per ml.

### Human epithelial cells cultured *in vitro*

Sterile urine was collected from patient B and an age- and gender-matched control. The outgrowth epithelial cells from urine were collected and seeded onto a gelatin-coated 12-well plate in renal epithelial cell growth medium (REGM BulletKit, Lonza). When epithelial cells were 70%–80% confluent, all cells were subcultured at the next passage with a 1:4 split ratio. During passages 1–3, the epithelial cells were used for the following experiment or frozen in liquid nitrogen for future use. The genotype of cell lines was confirmed by PCR amplification and Sanger sequencing using the primer mentioned above.

### Statistical analysis

In the genetic association analysis, two control groups were used, namely, the public Chinese control group (Huabiao-5000 human-exome-sequencing-data of general Chinese-Han-population, https://www.biosino.org/wepd) and the EAS_gnomAD control group (*CEP192* variants data downloaded from http://www.gnomad-sg.org/). Logistic regression analysis was used to estimate the association between a variant (that appears twice or more in 1264 infertile males) and infertility. Statistical significance was considered when p < 0.05. Statistical analyses were conducted using IBM SPSS 20.0 (IBM SPSS, Inc., Chicago, IL).

