## supplementary methods, tabs and figs for "Mosaic variegated aneuploidy syndrome with tetraploid, and predisposition to male infertility triggered by mutant *CEP192*"

### Clinical Details of *CEP192* mutation positive individuals with interests

### Patient B

Pateint B showed intrauterine growth retardation, premature delivery. Her birth weight was -3.45SD (according to WHO Anthro3.0 procedures). She was evaluated in our clinic with the following phenotypes were observed: a) low weight, her weight was <-3SD, according to 2005 nine provinces/cities children physical development survey data in China); b) low height, her height was <-3SD; c) microcephaly, her head circumference was <-3SD; d) hyper-pigmentation evenly prevailed the body); e) special facial features similar to MVA syndrome, including moderate ptosis, beak-like nose, micrognathia and frontal bossing; f) limb extremities abnormalities, including brachydactyly, syndactyly and epiphyseal plates were absent in some bones. Results of two rounds of GTG-banding was provided (Supplementary Table 14). The frequency of PSCS was analyzed in 300 metaphase cells. A total of 21 cells (7%) with PSCS was identified.

### Patient A

Patient A phenotypes were similar to that of Patient B. He showed intrauterine growth retardation, premature delivery, birth weight was -4.73SD. He was evaluated in our clinic. The following phenotypes were observed: a) low weight, her weight was <-3SD; b) low height, her height was <-3SD; c) microcephaly, her head circumference was <-3SD; d) hyper-pigmentation evenly prevailed the body; e) special facial features, including moderate ptosis, beak-like nose, micrognathia and frontal bossing; f) limb extremities abnormalities, including brachydactyly, syndactyly, epiphyseal plate was absent in some bones; g) his testes were smaller than normal (Supplementary Table 1). Results of GTG-banding was provided (Supplementary Table 15). The frequency of PSCS was analyzed in 300 metaphase cells. A total of 37 cells (12%) with PSCS was identified.

### Patient C

His testes were smaller than normal (Supplementary Table 1). Semen analyses conducted for him showed normal semen volume but the number of sperm was reduced (Supplementary Table 1). He has been married for 19 years, without contraception, the couple has had two pregnancies, suggesting the status of the reduced fertility. Results of two rounds of GTG-banding was provided

(Supplementary Table 16). The frequency of PSCS was analyzed in 300 metaphase cells. A total of 9 cells (3%) with PSCS was identified.

**Patient D**

He was complete infertile. By physical examination, his testes were smaller than normal (Supplementary Table 1). At least two semen analyses were conducted on him. Results showed normal semen volume but almost no sperm was obtained (Supplementary Table 1). Testicular biopsy performed in him disclosed the change of vacuolar degeneration, and no mature sperm was seen, in all available seminiferous tubules of of patient D(Figure 1B). In addition,

the germ cell number was reduced obviously in seminiferous tubules of p

atient D (Figure 1B). GTG-banding was performed at local hospital. The MVA and tetraploidy cells can be seen on his slides (Supplementary Table 17). The frequency of PSCS was analyzed in 300 metaphase cells. A total of 14 cells (5%) with PSCS was identified.

**Patient E**

He was a patient with primary infertility. Routine semen analyses revealed oligoasthenozoospermia with normal volume (Supplementary Table 1). His hormone levels were normal (Supplementary Table 1). B ultrasonography showed that his bilateral testicular size is smaller than normal (Supplementary Table 1). Results of GTG-banding was provided (Supplementary Table 18). The frequency of PSCS was analyzed in 300 metaphase cells. A total of 4 cells (1%) with PSCS was identified.

**Patient F**

He was a patient with oligoasthenozoospermia (Supplementary Table 1). His wife failed to conceive after more than six years of marriage, despite unprotected sexual intercourse with ejaculation. His bilateral testes were smaller than normal (Supplementary Table 1). Cytogenetic analysis was not performed on him.

**Patient G**

He was infertile. His wife failed to conceive after more than two years of marriage, despite unprotected sexual intercourse with ejaculation. His hormone levels were in normal (Supplementary Table 1). His bilateral testes were small (Supplementary Table 1). Semen analyses showed oligoasthenozoospermia with normal volume (Supplementary Table 1). Results of GTG-banding was provided (Supplementary Table 19). The frequency of PSCS was analyzed in 96 metaphase cells. A total of 13 cells (14%) with PSCS was identified.

**Supplementary Table 1:** Andrology-data for *CEP192*-mutant-individuals with infertility.

| | Ref values | Patient C | Patient D | Patient A | Patient E | Patient F | Patient G |
|---|---|---|---|---|---|---|---|
| ***CEP192* variants** | WT/WT | WT/c.5750A>G | WT/c.1912C>T | c.5750A>G/ c.1912C>T | WT/c.5750A>G | WT/c.5750A>G | WT/c.5750A>G |
| **Semen volume (mL)[a]** | ＞1.5 | 2.8 | 2.4 | NA | 2 | 2.1 | 2.71 |
| **Sperm Concentrationa (millions/mL) [a]** | ＞15 | 0.74 | 0.0019 | NA | 1.71 | 2.84 | 1.31 |
| **Motile sperm (%)[a]** | ＞40 | 1 | 0 | NA | 5.56 | 3.33 | 4.20 |
| **Testosterone (ng/mL) [b]** | 2.49-8.36 | 3.38 | 7.94 | 4.12 | 5.05 | NA | NA |
| **FSH (mIU/mL) [b]** | 1.5-12.4 | 3.5 | 19.7 | 14.5 | 7.04 | NA | NA |
| **LH (mIU/mL) [b]** | 1.7-8.6 | 2.2 | 8.4 | 6 | 2.21 | NA | NA |
| **Prolactin (ng/mL) [b]** | 4.04-15.2 | 8.7 | 19.8 | 12.7 | NA | NA | NA |
| **Testicular volume (mL) [c]** | 12-20 mL | L 9.6 | L 6.3 | L 4.5 | L 5.7 | L 10 | L 7.1 |
| | | R 12.1 | R 5.8 | R 4.1 | R 4.9 | R 10 | R 5.3 |

NA: data not available; Ref, reference; WT, wild type allele.

a: Routine semen analyses were performed at least twice. Reference values were published by WHO in 2010 (WHO laboratory manual for the examination and processing of human semen).

b: Reference values were suggested by the local clinical laboratory. FSH, follicle-stimulating hormone; LH, luteinizing hormone.

c: Testicular volume was calculated by ultrasound measurement (formula: length x width x height x 0.52) (PMID: 17270639); L, left; R, right. Ref value was according to *ANDROLOGY* (in Chinese, People's Medical Publishing House, Edited by Yinglu Guo and Liquan Hu).

Note: the II-1, II-5 and III-1 were from the index family; Y8147,MD3337 and T00634 were from 1264 males with idiopathic infertility from general population.

**Supplementary Figure 1:** Representative GTG-banding-primary figures for patient A (left) and patient B (right) that correspondent to Figure 2

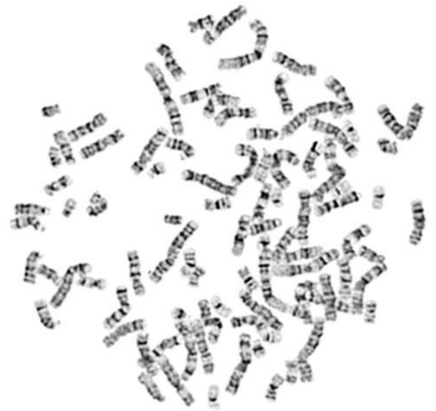

**92, XXYY**

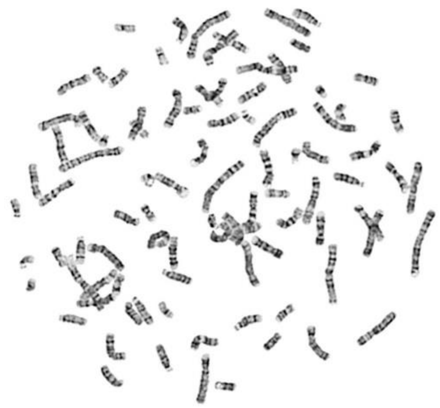

**91, XXXX, -11**

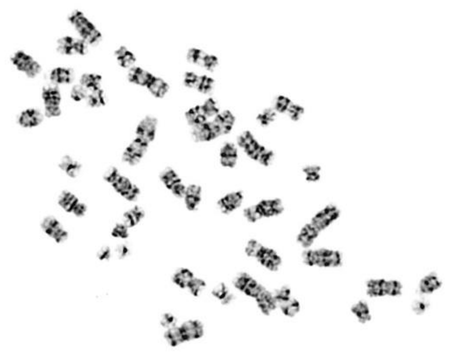

**46, XY, +18, -21**

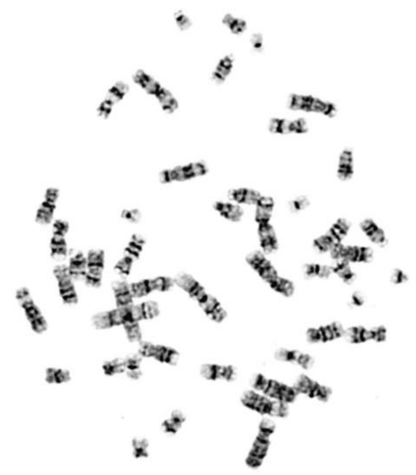

**48, XX, +8, +10**

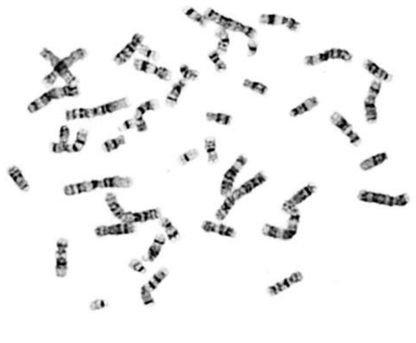

**47, XY, +18**

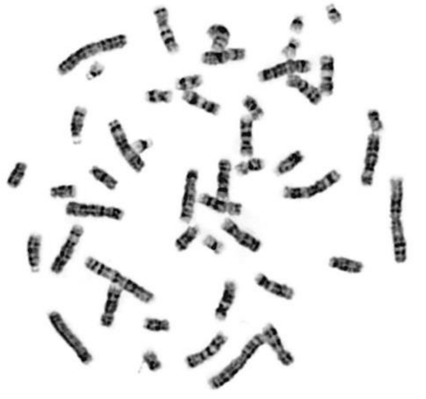

**48, XX,+8, +18**

**Supplementary Table 2:** Quality data of exome sequencing for the index family.

| Sample | Parent | Parent | Patient A | Patient B |
|---|---|---|---|---|
| Clean reads (Read Count) | 70941634 | 75692830 | 69030578 | 72342574 |
| Mapped reads (Read Count) | 70808731 (99.81%) | 75566116 (99.83%) | 68855792 (99.75%) | 72189146 (99.79%) |
| Fraction of effective bases on target | 68.30% | 65.60% | 65.00% | 65.90% |
| Fraction of effective bases on or near target | 87.30% | 86.70% | 87.30% | 87.20% |
| Average sequencing depth on target | 119.25 | 122.11 | 110.12 | 117.09 |
| Coverage of target region | 99.60% | 99.60% | 99.90% | 99.90% |
| Fraction of target covered at least 4x | 99.60% | 99.60% | 99.80% | 99.80% |
| Fraction of target covered at least 10x | 99.30% | 99.30% | 99.50% | 99.50% |
| Fraction of target covered at least 20x | 98.20% | 98.50% | 98.60% | 98.60% |
| Fraction of target covered at least 50x | 85.60% | 87.30% | 87.30% | 89.70% |
| Fraction of target covered at least 100x | 49.00% | 50.90% | 47.00% | 53.10% |

**Supplementary Figure 2:** The analysis strategy to search for candidate gene for the index family.

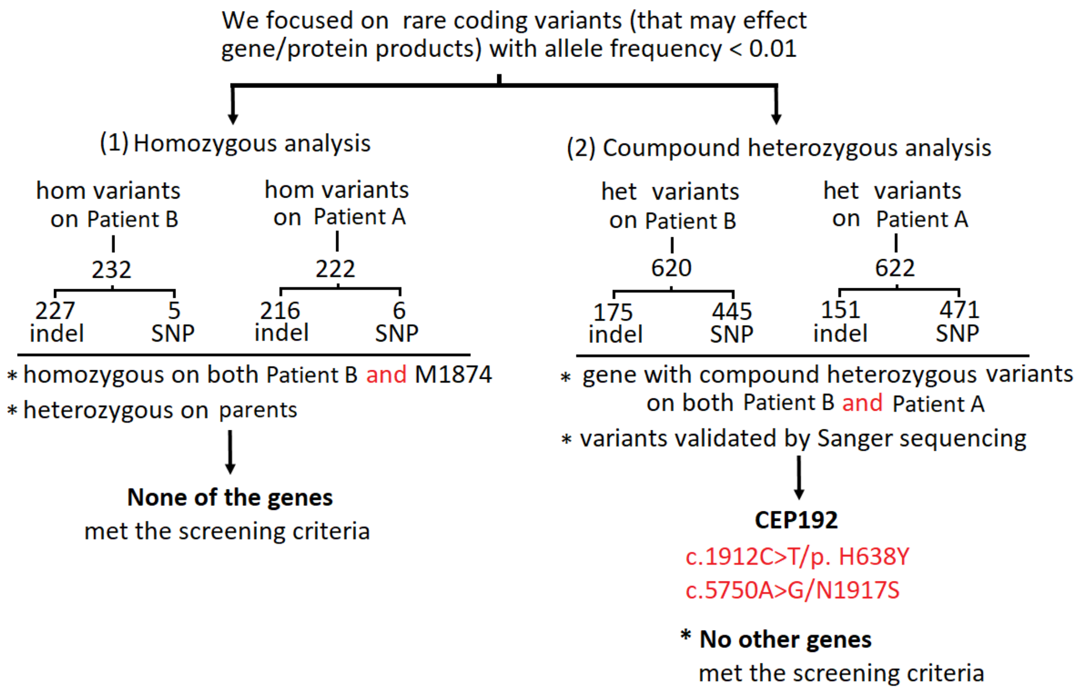

**Supplementary Table 3:** Several *CEP192* rare variants (MaF<0.001 in Eas_gnomAD v2.1.1) significantly associated with infertility in 1264 infertile males. Control= Public Huabiao-human-exome-sequencing-data of 5000 general Chinese-Han-population (https://www.biosino.org/wepd; PMID: 34416338).

| | p.1416 | | p.1394 | | p.393 | | p.47 | | p.1917 | |
|---|---|---|---|---|---|---|---|---|---|---|
| | N1416N | wide-type | L1394V | wide-type | G393E | wide-type | R47R | wide-type | N1917S | wide-type |
| Patient | 8 | 2520 | 5 | 2523 | 5 | 2523 | 5 | 2523 | 3 | 2525 |
| Control | 2 | 9916 | 4 | 9914 | 2 | 9920 | 2 | 9920 | 0 | 9918 |
| P | <0.001 | | 0.009 | | 0.001 | | 0.001 | | 0.001 | |
| OR(95% CI) | 15.740 (3.340, 74.165) | | 4.912 (1.318, 18.305) | | 9.830 (1.906, 50.694) | | 9.830 (1.906, 50.694) | | 15.707(1.755, 140.590) | |

CI, confidence interval

**Supplementary Table 4:** Statistic analysis for several *CEP192* rare variants with interests in 1264 infertile males and controls. Controls= Eas_gnomAD v2.1.1 on Public database (http://www.gnomad-sg.org/)

| | p.1416 | | p.1394 | | p.393 | | p.47 | | p.1917 | |
|---|---|---|---|---|---|---|---|---|---|---|
| | N1416N | wide-type | L1394V | wide-type | G393E | wide-type | R47R | wide-type | N1917S | wide-type |
| Patient | 8 | 2520 | 5 | 2523 | 5 | 2523 | 5 | 2523 | 3 | 2525 |
| Control | 2 | 19952 | 16 | 19936 | 2 | 10898 | 1 | 1555 | 2 | 18392 |
| P | <0.001 | | 0.068 | | <0.001 | | 0.279 | | 0.015 | |
| OR(95% CI) | 31.667(6.721, 149.203) | | 2.469 (0.904, 6.746) | | 10.799 (2.094, 55.691) | | 3.082 (0.360, 26.402) | | 10.926 (1.825, 65.419) | |

**Supplementary Table 5:** Testes size of infertile males with the enriched-*CEP192* variants. **Note:** the reference-testicular size for Chinese was 12-15mL.

| **Samples** | **Variants** | **Testis size/mL (left; right)** |
|---|---|---|
| Patient E | c.5750A>G: p.N1917S | 5.7; 4.9 |
| Patient F | | 10; 10 |
| Patient G | | 7.1; 5.3 |
| Patient 1 | c.4248T>C: p.N1416N | 6.1； 6.1 |
| Patient 2 | | 3； 4 |
| Patient 3 | | 10.1; 9.1 |
| Patient 4 | | 11.3; 11.7 |
| Patient 5 | | 15; 15 |
| Patient 6 | | 8.6; 8.2 |
| Patient 7 | | 9.2; 11.5 |
| Patient 8 | | 11.2; 11.5 |
| Patient 9 | c.4180C>G: p.L1394V | 4.8; 4 |
| Patient 10 | | 7.4; 6.4 |
| Patient 11 | | 12; 12 |
| Patient 12 | | 12;12 |
| Patient 13 | | 4.5; 4.4 |
| Patient 14 | c.1178G>A: p.G393E | NA |
| Patient 15 | | 4.8; 3.9 |
| Patient 16 | | 15; 18 |
| Patient 17 | | 17.4; 13.9 |
| Patient 18 | | 15; 16.7 |
| Patient 19 | c.141G>A: p.R47R | 11.9; 13.1 |
| Patient 20 | | NA |
| Patient 21 | | 4.8; 5.0 |
| Patient 22 | | 8; 10 |
| Patient 23 | | 13; 9.5 |

**Supplementary Figure 3:** Strategy for generation of a C57BL/6N mouse model with knock out (KO) *Cep192* by CRISPR/Cas-mediated genome engineering. **A:** Schematic depiction of targeting strategy. Genomic region of mouse *Cep192* locus is diagrammed below (gene is oriented from left to right). 46 exons are identified. Exon 6-41 will be selected as target site. Cas9 and gRNA will be co-injected into fertilized eggs for KO mice production. Note: i) Exon 6 starts from about 5.87% of the coding region of mice *Cep192* gene; ii) Exon 6-41 covers 86.66% of the coding region; iii) The size of effective KO region: 61482 bp and the KO region does not have any other known gene; **B:** gRNA target sequence for generation of the *Cep192* KO mice; **C:** A representative Sanger sequencing figure of the knock-out mice; **D:** Primers and PCR conditions of the assay of the CRISPR-induced knock-out mice.

**A:**

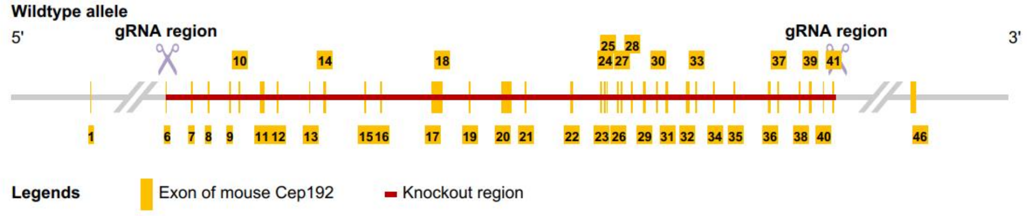


**B:** **gRNA target sequence**

gRNA1 (matching reverse strand of gene):
GGCGCTGGGCCTCTTTAAAGTGG
gRNA2 (matching reverse strand of gene):
GGAGACAAACTCAAGTGACGAGG

**C:**

**Positive animals (Deleted 62487 bp)**

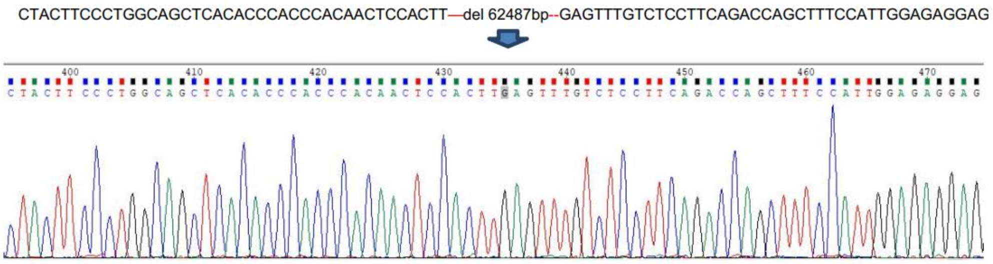


**D:**

**Inter-cross heterozygous targeted mice to generate homozygous targeted mice:**
PCR Primers1 (Annealing Temperature 60.0 ºC):
F1: 5'-GCTGTCAAGAAAGTGACAAAACAACC-3'
R1: 5'-GTAGACAGCACTACTGTTGATGAG-3'
Product size: 622 bp
PCR Primers2 (Annealing Temperature 60.0 ºC):
F2: 5'-CGGATATTTGGGGCAAACAGTCAA-3'
R1: 5'-GTAGACAGCACTACTGTTGATGAG-3'
Product size: 430 bp

**Homozygous:** one band with 622 bp
**Heterozygous:** two bands with 622 bp and 430 bp
**WT:** one band with 430 bp

**Supplementary Figure 4:** Strategy for generation of a C57BL/6N mouse model with missense variant at mouse *Cep192* locus by CRISPR/Cas-mediated genome engineering. **A:** Schematic depiction of targeting strategy. Genomic region of mouse *Cep192* locus is diagrammed below (gene is oriented from left to right; E32=exon32). Solid bars represent ORF; open bars represent UTRs; **B:** gRNA target sequence, Links of gRNA vectors on Vector Builder gRNA1: https://en.vectorbuilder.com/vector/VB190818-1081fuu.html, gRNA2: https://en.vectorbuilder.com/vector/VB190818-1082eup.html; **C:** Assay of CRISPR-induced mutation. The target region of mouse *Cep192* locus will be amplified by PCR with specific primers. PCR product will be sequenced to confirm targeting; **D:** Representative picture of Sanger sequencing profile of the mutant mice. **Note:** mice N1892 amino acid is equals to human N1917 amino acid.

**A:**

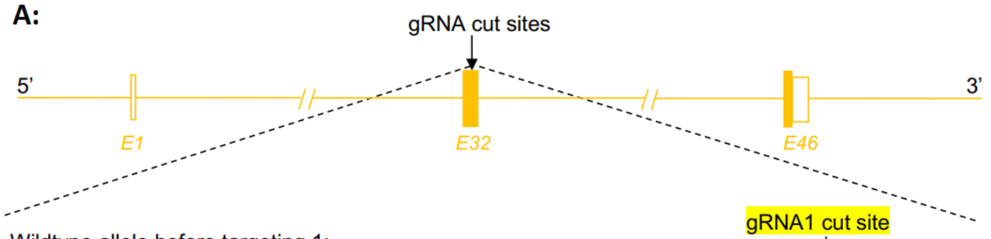


Wildtype allele before targeting 1:

TTGATAACAATGAATGACTTAATTCCTGGCAAAGAAAGTAAAGTTGTTTTTTCTATTCGC(AAC)ACTGGCTCCAG
AGCAGCCTTTGTTAGAGCAGTATGTTTTAAAGAGTCTCAAAGAAAAGTT

Mutant allele after targeting 1:

TTGATAACAATGAATGACTTAATTCCTGGCAAAGAAAGTAAAGTTGTTTTTTCTATTCGC(AGC)ACTGGCTCCAG
AGCAGCCTTTGTTAGAGCAGTATGTTTTAAAGAGTCTCAAAGAAAAGTT

**B:**

**gRNA target sequence**
**gRNA1** (matches forward strand of gene):
GTTTTTTCTATTCGCAACACTGG (priority)
**gRNA2** (matches reverse strand of gene):
TGCTCTAACAAAGGCTGCTCTGG

**C:**

**Assay of CRISPR-induced mutation**
Primer sequence:
Mouse Cep192 (N1892S)-F:
CTAGCTCAAGGATGCAAACACTAGAAAC
Mouse Cep192 (N1892S)-R:
ACAGCAGCTCTCCCAGAAAGACTAG
**Expected PCR product size:**
Wildtype allele: 521 bp
Mutant allele: 521 bp

**D:**

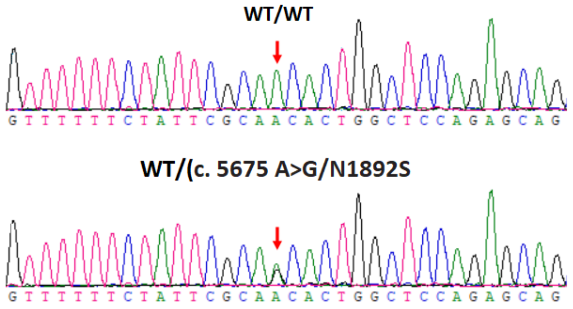

**Supplementary Table 6:** Quantification of progeny from crosses between *Cep192*$^{+/-}$ mice

| Mouse Pairs | | Age (days) | Mating from | Persistent Mating for 210 days | | | | | | Giving Birth |
|---|---|---|---|---|---|---|---|---|---|---|
| | | | | Litter 1 | Litter 2 | Litter 3 | Litter 4 | Litter 5 | Litter 6 | |
| 1 | ♂ KO-HET-D1-L1 | 55d | 20.09.07 | 20.09.29 | 20.11.18 | 21.01.17 | 21.02.23 | 21.04.08 | N | WT:7 HET:4 HOM:0 |
| | ♀ KO-HET-B2-L2 | 78d | | **5** | **2** | **2** | **1** | **1** | | |
| 2 | ♂ KO-HET-A1 | 164d | 20.09.07 | 20.10.01 | 20.11.06 | 21.01.04 | N | | | WT:6 HET:11 HOM:0 |
| | ♀ KO-HET-B8 | 78d | | **8** | **6** | **3** | | | | |
| 3 | ♂ KO-HET-D7-R3 | 55d | 20.09.07 | 20.10.08 | 20.10.28 | 20.12.05 | N | | | WT:5 HET:12 HOM:0 |
| | ♀ KO-HET-B6-R2 | 78d | | **4** | **4** | **9** | | | | |
| 4 | ♂ KO-HET-D5-R1 | 55d | 20.09.07 | 20.10.13 | 20.11.20 | 21.02.14 | 21.03.15 | N | | WT:10 HET:10 HOM:0 |
| | ♀ KO-HET-B3-L3 | 78d | | **7** | **8** | **3** | **2** | | | |
| 5 | ♂ KO-HET-D4-L4 | 55d | 20.09.07 | 20.11.11 | N | | | | | WT:0 HET:3 HOM:0 |
| | ♀ KO-HET-B7-R3 | 78d | | **3** | | | | | | |
| 6 | ♂ KO-HET-AC6-R2 | 101d | 21.04.16 | 21.05.12 | N | | | | | WT:0 HET:5 HOM:0 |
| | ♀ KO-HET-AH9-FL1 | 59d | | **5** | | | | | | |
| 7 | ♂ KO-HET-AG3-L3 | 68d | 21.04.16 | 21.05.17 | 21.07.05 | 21.08.22 | N | | | WT:2 HET:6 HOM:0 |
| | ♀ KO-HET-AB1-L1 | 58d | | **5** | **2** | **1** | | | | |
| 8 | ♂ KO-HET-AH1-L1 | 60d | 21.04.16 | N | | | | | | WT:0 HET:0 HOM:0 |
| | ♀ KO-HET-AD2-L2 | 78d | | | | | | | | |
| 9 | ♂ KO-HET-AF3-L3 | 69d | 21.04.16 | N | | | | | | WT:0 HET:0 HOM:0 |
| | ♀ KO-HET-AD2-L2 | 70d | | | | | | | | |
| 10 | ♂ KO-HET-I5-R1 | 52d | 21.11.05 | 21.11.24 | 21.12.19 | 22.02.01 | 22.04.01 | 22.04.25 | 22.06.08 | WT:11 HET:15 HOM:0 |
| | ♀ KO-HET-G8-R4 | 81d | | **7** | **6** | **5** | **3** | **3** | **2** | |
| 11 | ♂ KO-HET-E4-L4 | 103d | 21.11.05 | 21.11.30 | 22.01.07 | 22.03.30 | 22.05.03 | N | | WT:5 HET:4 HOM:0 |
| | ♀ KO-HET-D8-R4 | 115d | | **1** | **2** | **5** | **1** | | | |
| 12 | ♂ KO-HET-I2-L2 | 52d | 21.11.05 | 21.11.25 | 21.12.20 | 22.02.06 | 22.04.07 | N | | WT:6 HET:9 HOM:0 |
| | ♀ KO-HET-E3-L3 | 103d | | **1** | **6** | **4** | **4** | | | |
| | | | | | | | | | | **Total: WT:52 HET:79 HOM:0** |

**Supplementary Table 7:** Quantification of progeny from crosses between *Cep192*$^{+/M}$ mice

| Mouse Pairs | | Age (days) | Mating from | Persistent Mating for 210 days | | | | | | Giving Birth |
|---|---|---|---|---|---|---|---|---|---|---|
| | | | | **Litter 1** | **Litter 2** | **Litter 3** | **Litter 4** | **Litter 5** | **Litter 6** | |
| 1 | ♂ CEP192+/- -A3-L3 | 50d | 21.02.07 | 21.03.01 | 21.04.22 | 21.05.31 | 21.07.13 | 21.08.31 | 21.09.26 | WT: 10<br>HET: 20<br>HOM: 0 |
| | ♀ CEP192+/- -B7-R3 | 57d | | **7** | **6** | **7** | **5** | **3** | **2** | |
| 2 | ♂ CEP192+/- -B8-R4 | 50d | 21.02.07 | 21.03.02 | 21.04.10 | 21.06.06 | 21.07.25 | N | | WT: 4<br>HET: 11<br>HOM: 0 |
| | ♀ CEP192+/- -A2-L2 | 57d | | **4** | **7** | **3** | **1** | | | |
| 3 | ♂ CEP192+/- -B2-L2 | 50d | 21.02.07 | 21.03.02 | 21.04.15 | N | | | | WT: 4<br>HET: 10<br>HOM: 0 |
| | ♀ CEP192+/- -A4-L4 | 57d | | **5** | **9** | | | | | |
| 4 | ♂ CEP192+/- -F1-L1 | 74d | 21.03.24 | 21.05.17 | N | | | | | WT: 6<br>HET: 3<br>HOM: 0 |
| | ♀ CEP192+/- -L3-L3 | 69d | | **9** | | | | | | |
| 5 | ♀ CEP192+/- -F4-L4 | 74d | 21.03.24 | 21.04.24 | 21.06.11 | N | | | | WT: 3<br>HET: 6<br>HOM: 0 |
| | ♂ CEP192+/- -H9-FL1 | 61d | | **5** | **4** | | | | | |
| 6 | ♂ CEP192+/- -F3-L3 | 74d | 21.03.24 | 21.04.24 | 21.05.16 | 21.06.21 | 21.07.19 | 21.08.19 | N | WT: 13<br>HET: 8<br>HOM: 0 |
| | ♀ CEP192+/- -H11-FL3 | 61d | | **8** | **7** | **2** | **1** | **3** | | |
| 7 | ♀ CEP192+/- -F6-R2 | 55d | 21.03.24 | 21.04.27 | N | | | | | WT: 0<br>HET: 2<br>HOM: 0 |
| | ♂ CEP192+/- -H10-FL2 | 61d | | **2** | | | | | | |
| 8 | ♀ CEP192+/- -F2-L2 | 55d | 21.03.24 | 21.04.26 | 21.05.24 | 21.07.01 | 21.07.24 | 21.09.25 | 21.11.21 | WT: 7<br>HET: 10<br>HOM: 0 |
| | ♂ CEP192+/- -H12-FL4 | 61d | | **6** | **3** | **2** | **4** | **1** | **1** | |
| 9 | ♂ CEP192+/- -Y5-R1 | 121d | 21.07.28 | 21.08.29 | 21.10.14 | 21.12.02 | 22.01.05 | N | | WT: 6<br>HET: 7<br>HOM: 0 |
| | ♀ CEP192+/- -AA3-L3 | 94d | | **3** | **6** | **3** | **1** | | | |
| 10 | ♂ CEP192+/- -Y4-L4 | 103d | 21.07.28 | 21.09.17 | N | | | | | WT: 0<br>HET: 2<br>HOM: 0 |
| | ♀ CEP192+/- -AA1-1L | 94d | | **2** | | | | | | |
| 11 | ♂ CEP192+/- -R7-R3 | 103d | 21.07.28 | 21.10.06 | N | | | | | WT: 2<br>HET: 3<br>HOM: 0 |
| | ♀ CEP192+/- -AA5-R1 | 94d | | **5** | | | | | | |
| 12 | ♂ CEP192+/- -R9-FL1 | 103d | 21.07.28 | 21.09.01 | 21.10.05 | 21.10.26 | 21.11.20 | 22.01.09 | 22.02.13 | WT: 9<br>HET: 12<br>HOM: 0 |
| | ♀ CEP192+/- -AA2-L2 | 94d | | **8** | **2** | **2** | **4** | **3** | **2** | |
| | | | | | | | | | | **Total**<br>WT: 64<br>HET: 94<br>HOM: 0 |

**Supplementary Table 8:** Quantification of progeny from crosses between *Cep192*$^{+/-}$ (KO) and *Cep192*$^{+/M}$ (KI) mice.

| | Mouse Pairs | Age (days) | Mating from | Persistent Mating for 210 days | | | | | Giving Birth |
|---|---|---|---|---|---|---|---|---|---|
| | | | | **Litter 1** | **Litter 2** | **Litter 3** | **Litter 4** | **Litter 5** | |
| 1 | ♂ CEP192+/- -FL3-R2 | 61d | 20.12.11 | 21.01.09 | 21.03.22 | N | | | WT: 3<br>CEP192+/- : 7<br>KO-HET: 3<br>KO/KI: 0 |
| | ♀ KO-HET-J2-L2 | 73d | | **7** | **6** | | | | |
| 2 | ♂ CEP192+/- -LL3-R1 | 61d | 20.12.11 | N | | | | | WT: 0<br>CEP192+/- : 0<br>KO-HET: 0<br>KO/KI: 0 |
| | ♀ KO-HET-J4-L4 | 73d | | | | | | | |
| 3 | ♂ CEP192+/- -FL+R1 | 61d | 20.12.11 | 21.01.13 | 21.02.27 | 21.05.28 | N | | WT: 6<br>CEP192+/- : 5<br>KO-HET: 8<br>KO/KI: 0 |
| | ♀ KO-HET-M1-L1 | 59d | | **9** | **7** | **3** | | | |
| 4 | ♂ KO-HET-H5 | 162d | 20.12.11 | 21.01.17 | 21.03.15 | N | | | WT: 4<br>CEP192+/- : 0<br>KO-HET: 1<br>KO/KI: 0 |
| | ♀ CEP192+/- -FL3-L4 | 61d | | **3** | **2** | | | | |
| 5 | ♂ KO-HET-K5-R1 | 68d | 21.03.10 | 21.04.01 | 21.05.07 | 21.06.10. | 21.08.07 | 21.09.27 | WT: 5<br>CEP192+/- : 6<br>KO-HET: 5<br>KO/KI: 0 |
| | ♀ CEP192+/- -E5-R1 | 61d | | **5** | **3** | **4** | **2** | **2** | |
| 6 | ♂ KO-HET-K7-R3 | 68d | 21.03.10 | 21.04.04 | 21.06.04 | 21.09.10 | N | | WT: 4<br>CEP192+/- : 7<br>KO-HET: 4<br>KO/KI: 0 |
| | ♀ CEP192+/- -E10-FL2 | 61d | | **8** | **4** | **3** | | | |
| 7 | ♂ KO-HET-Y2-L2 | 107d | 21.03.22 | 21.04.17 | 21.06.11 | 21.08.22 | N | | WT: 7<br>CEP192+/- : 4<br>KO-HET: 4<br>KO/KI: 0 |
| | ♀ CEP192+/- -H3-L3 | 58d | | **6** | **6** | **3** | | | |
| 8 | ♂ KO-HET-Y6-R2 | 107d | 21.03.22 | 21.04.09 | 21.05.29 | 21.08.24 | N | | WT: 6<br>CEP192+/- : 3<br>KO-HET: 3<br>KO/KI: 0 |
| | ♀ CEP192+/- -H2-L2 | 58d | | **6** | **4** | **2** | | | |
| | | | | | | | | | **Total:**<br>WT: 35<br>CEP192+/- : 32<br>KO-HET: 28<br>KO/KI: 0 |

**Supplementary Figure 5:** Homozygous fertilized eggs can develop to blastocyst in vitro. In vitro fertilization was performed by using the sperms and eggs that came from the *Cep192*$^{+/-}$ C57BL/6N mice.

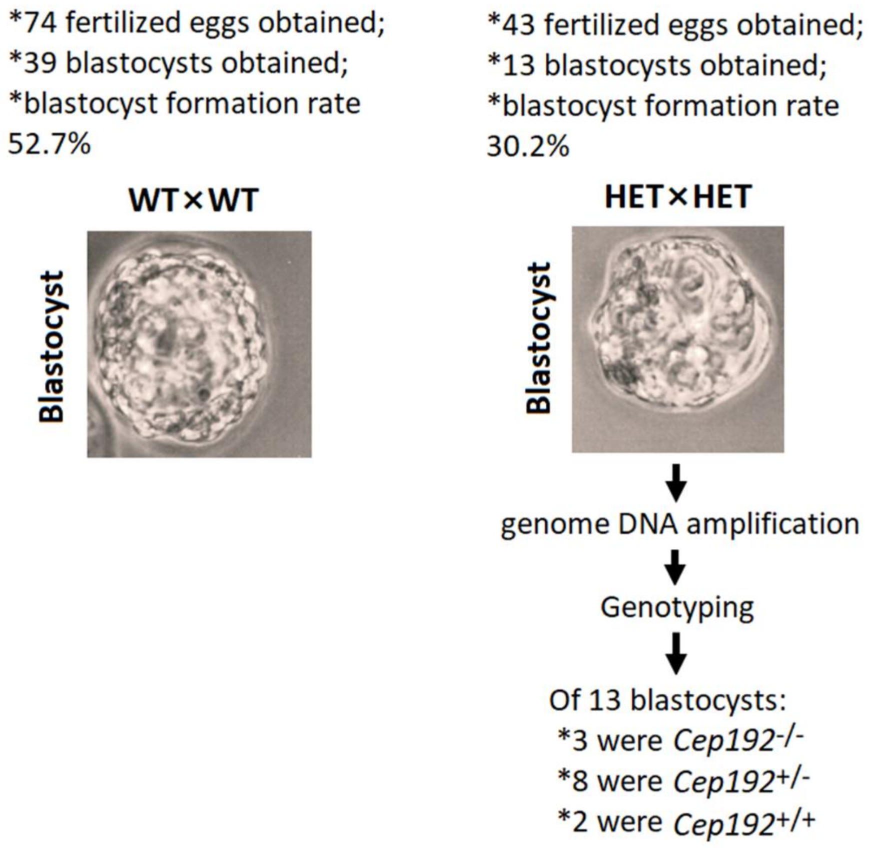

**Supplementary Figure 6:** Representative view of *Cep19*$^{-/-}$ embryos were alive to E7.5 and appeared morphologically normal. The *Cep192*$^{+/-}$ female mated to *Cep192*$^{+/-}$ male in day 1 (overnight), seeing vagina plug=0.5 day. On day 7.5, the pregnant female was dissected, and each embryo was isolated for genotyping. After genotyping, results showed that 3/11 were *Cep192*$^{-/-}$; 7/11 were *Cep192*$^{-/+}$ and 4/11 were *Cep192*$^{+/+}$. Specifically, in the box, we determined the top one was *Cep192*$^{+/+}$, and the bottom one is the *Cep192*$^{-/-,}$ no morphological difference can be identified externally.

**Note:** due to reduced fertility, we have intercrossed the mutant allele to Kunming mice for more embryos.

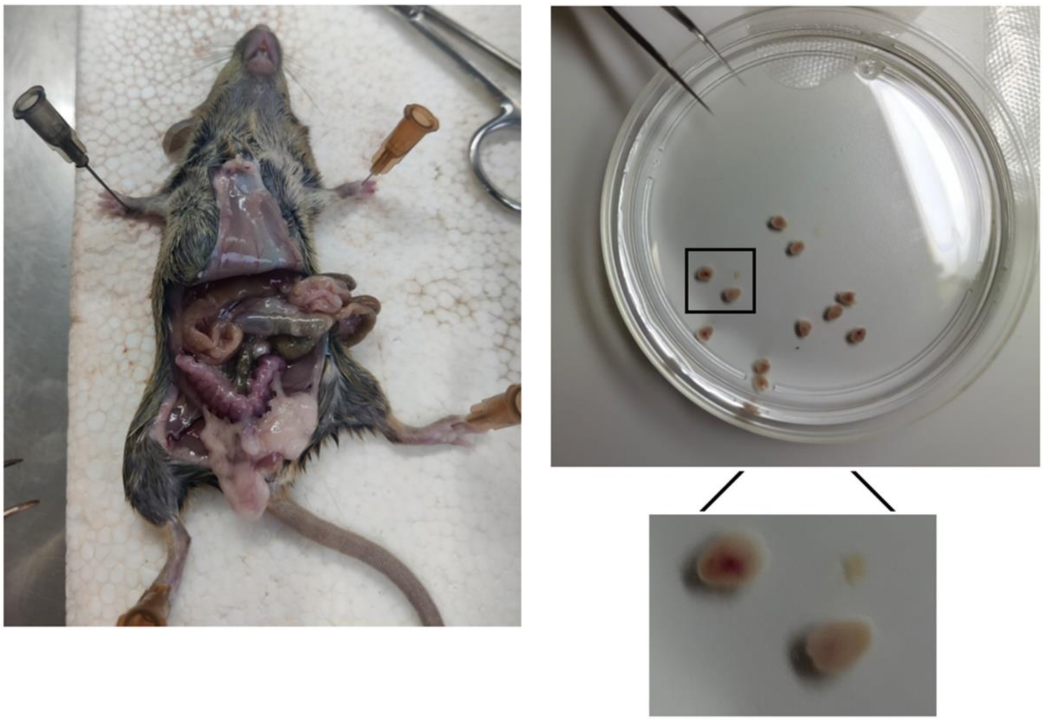

**Supplementary Figure 7:** Representative view of *Cep192*$^{-/-}$ embryos were dead in E11.5.

The *Cep192*$^{+/-}$ female mated to *Cep192*$^{+/-}$ male in day 1 (overnight), seeing vagina plug=0.5 day. On day 11.5, the pregnant female was dissected, and each embryo was isolated for genotyping and investigation

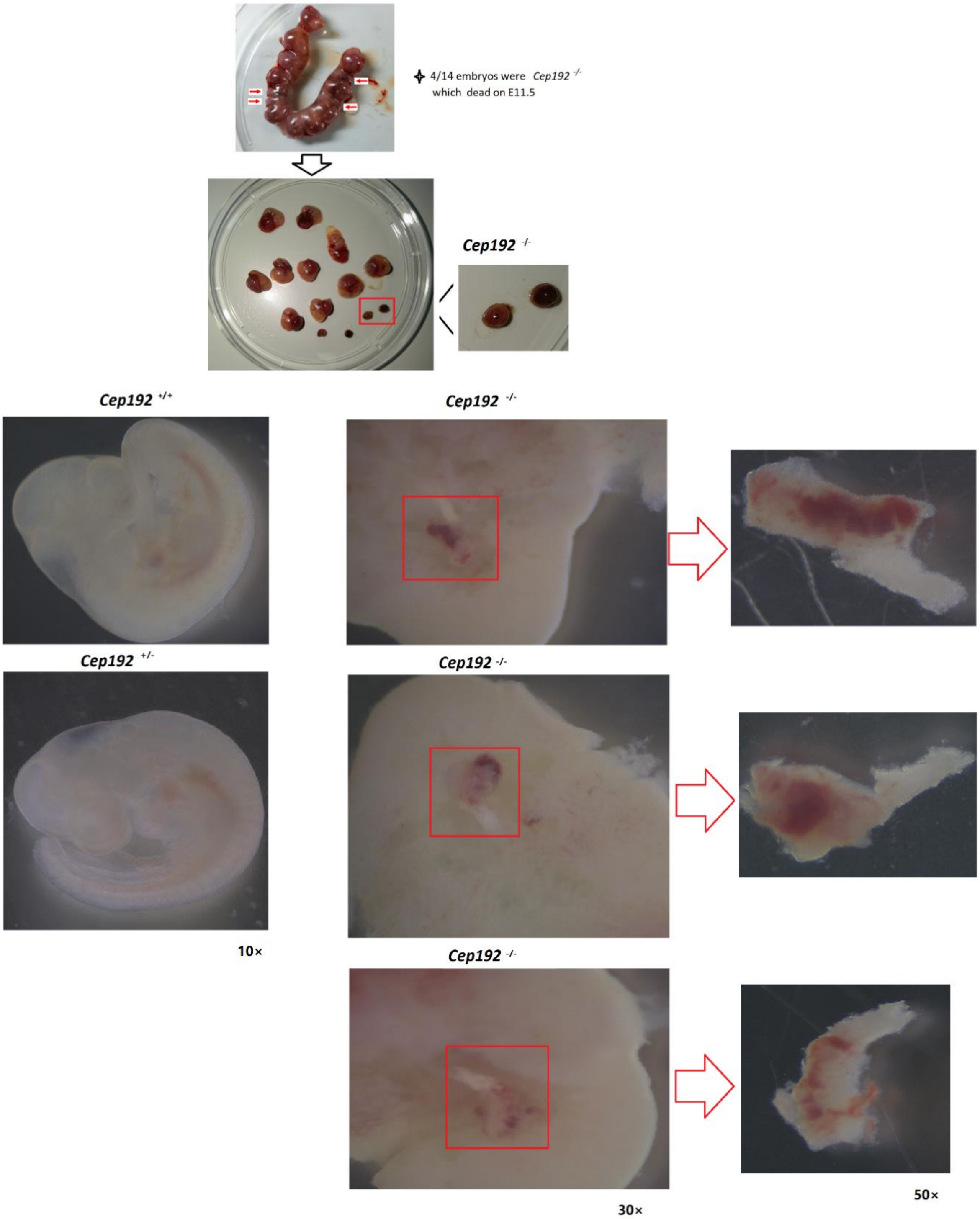

**Supplementary Figure 8:** Typical view of two pregnant females had several *Cep192*$^{+/-}$ embryos died in E19 and E14, respectively.

**Note:** The *Cep192*$^{+/+}$ female mated to *Cep192*$^{+/-}$ male in day 1 (overnight), seeing vagina plug=0.5day. On day 14 or 19, the pregnant female was dissected, and each embryo was isolated for genotyping.

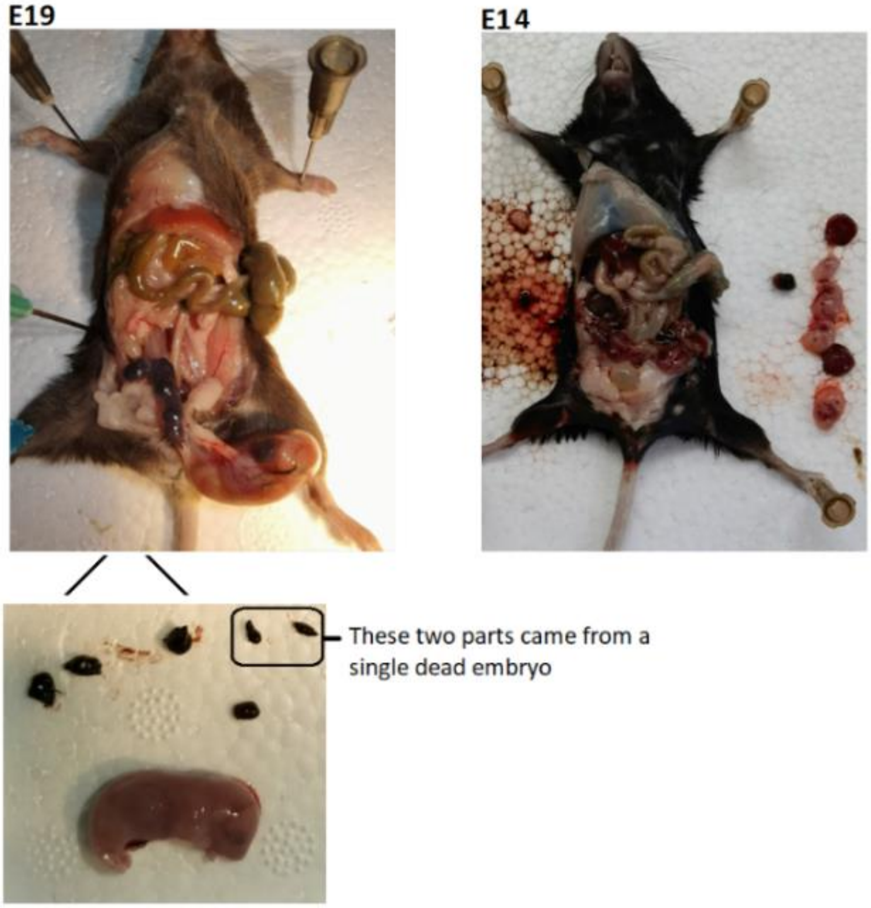

**Supplementary Figure 9:** A representative view of DAPI-staining, FISH and TUNNEL assays that performed on dead, *Cep192*$^{+-}$ embryos or normal *Cep192*$^{+/+}$ embryos.

**A:** Representative interphase nuclei monitored by FISH. **Left:** Cells from *Cep192*$^{+/+}$ embryos, three cell nuclei stained by two red (chromosome 15) and two green (chromosome X) signals; **Middle:** cells from *Cep192*$^{+/-}$ embryos (upper left, three red and five green signals; upper right: four green and four red signals; lower left: three green and two red signals; lower right: four green and three red signals); **Right:** diploidy cells, tetraploidy and MVA cell counts for *Cep192*$^{+/-}$ dead embryos or *Cep192*$^{+/+}$ normal embryos. For each genotype (*Cep192*$^{+-}$ or *Cep192*$^{+/+}$), a total of 100 cells were analyzed (n=3).

**B:** Cell apoptosis (cells stained by green) in cells from *Cep192*$^{+/+}$ normal cells or *Cep192*$^{+/-}$ stagnated embryos (n=3)

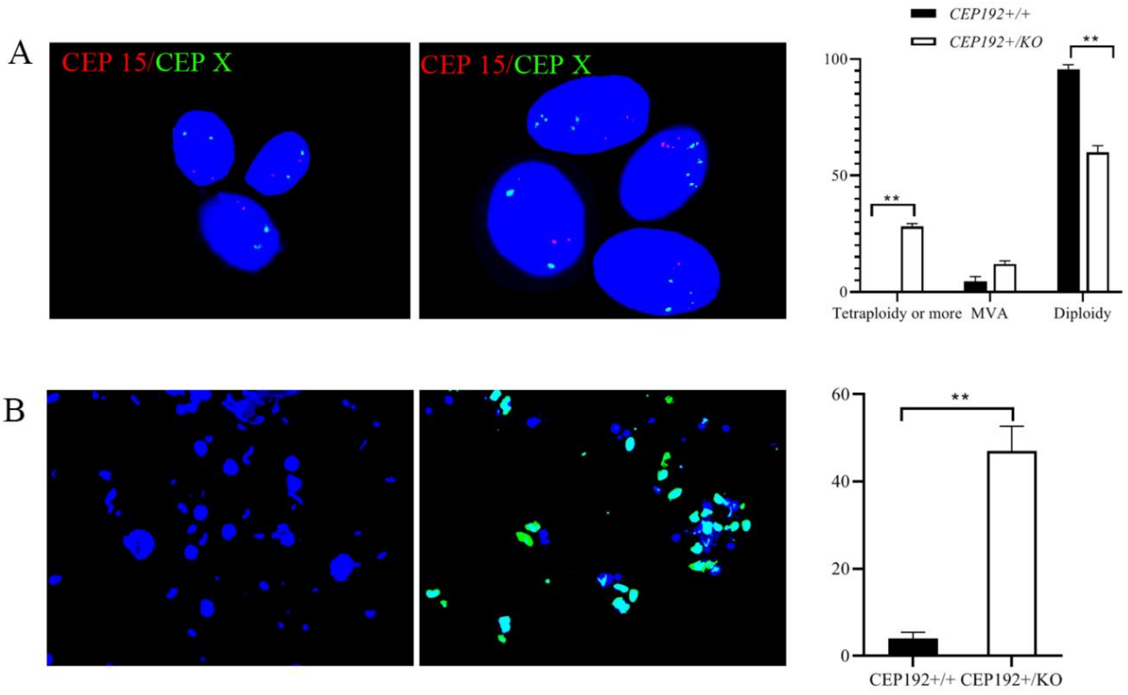

**Supplementary Table 9:** Reproduction of persistent mating *Cep192*$^{+/-}$ male mice with wild-type females for 210 days. *: days from mate to give the first birth; **: days from giving the first birth to the second birth; N: no birth

| Mouse Pairs | | Genotype | Age (days) | Persistent mating for 210 days | | | | | | | | | | | | | | | |
|---|---|---|---|---|---|---|---|---|---|---|---|---|---|---|---|---|---|---|---|
| | | | | Litter 1 days * | Litter 1 Pups | Mating days ** | Litter 2 Pups | Mating days | Litter 3 Pups | Mating days | Litter 4 Pups | Mating days | Litter 5 Pups | Mating days | Litter 6 Pups | Mating days | Litter 7 Pups | Overall Litters | Overall Pups |
| 1 | ♂AX7 | *Cep192*$^{+/-}$ | 59 | 30 | 7 | 43 | 6 | 53 | 3 | N | | | | | | | | 3 | 16 |
| | ♀AW3 | WT | 61 | | | | | | | | | | | | | | | | |
| 2 | ♂AW4 | *Cep192*$^{+/-}$ | 61 | 36 | 7 | 55 | 2 | N | | | | | | | | | | 2 | 9 |
| | ♀AS1 | WT | 62 | | | | | | | | | | | | | | | | |
| 3 | ♂ AN1 | *Cep192*$^{+/-}$ | 60 | 44 | 3 | 85 | 2 | N | | | | | | | | | | 2 | 5 |
| | ♀AX2 | WT | 59 | | | | | | | | | | | | | | | | |
| 4 | ♂AW5 | *Cep192*$^{+/-}$ | 61 | N | | | | | | | | | | | | | | 0 | 0 |
| | ♀AS4 | WT | 62 | | | | | | | | | | | | | | | | |
| 5 | ♂AT1 | *Cep192*$^{+/-}$ | 63 | N | | | | | | | | | | | | | | 0 | 0 |
| | ♀AO2 | WT | 59 | | | | | | | | | | | | | | | | |
| 6 | ♂AZ1 | *Cep192*$^{+/-}$ | 63 | 47 | 7 | 34 | 1 | N | | | | | | | | | | 2 | 8 |
| | ♀AT3 | WT | 63 | | | | | | | | | | | | | | | | |
| 7 | ♂BR2 | *Cep192*$^{+/-}$ | 60 | 36 | 6 | 70 | 5 | 66 | 5 | N | | | | | | | | 3 | 16 |
| | ♀BJ4 | WT | 63 | | | | | | | | | | | | | | | | |
| 8 | ♂BR3 | *Cep192*$^{+/-}$ | 60 | N | | | | | | | | | | | | | | 0 | 0 |
| | ♀BJ2 | WT | 59 | | | | | | | | | | | | | | | | |
| 9 | ♂BS1 | *Cep192*$^{+/-}$ | 63 | 21 | 7 | 21 | 10 | 24 | 5 | 29 | 4 | 41 | 1 | 37 | 2 | N | | 6 | 29 |
| | ♀IQ9 | WT | 59 | | | | | | | | | | | | | | | | |
| 10 | ♂BS2 | *Cep192*$^{+/-}$ | 63 | 21 | 8 | 43 | 7 | N | | | | | | | | | | 2 | 15 |
| | ♀BR7 | WT | 60 | | | | | | | | | | | | | | | | |

**Supplementary Table 10:** Reproduction of persistent mating *Cep192*$^{+/M}$ male (equivalent to human N1917S mutation) mice with wild-type females for 210 days. *: days from mate to give the first birth; **: days from giving the first birth to the second birth; N: no birth

| Mouse Pairs | | Genotype | Age (days) | Persistent mating for 210 days | | | | | | | | | | | | | | | |
|---|---|---|---|---|---|---|---|---|---|---|---|---|---|---|---|---|---|---|---|
| | | | | Mating days * | Litter 1 Pups | Mating days ** | Litter 2 Pups | Mating days | Litter 3 Pups | Mating days | Litter 4 Pups | Mating days | Litter 5 Pups | Mating days | Litter 6 Pups | Mating days | Litter 7 Pups | Overall Litters | Overall Pups |
| 1 | ♂AT5 | CEP192+/- | 63 | N | | | | | | | | | | | | | | 0 | 0 |
| | ♀AY1 | WT | 59 | | | | | | | | | | | | | | | | |
| 2 | ♂AT7 | CEP192+/- | 63 | 22 | 5 | 46 | 5 | N | | | | | | | | | | 2 | 10 |
| | ♀IJ4 | WT | 60 | | | | | | | | | | | | | | | | |
| 3 | ♂AX3 | CEP192+/- | 64 | 86 | 6 | N | | | | | | | | | | | | 1 | 6 |
| | ♀AZ6 | WT | 58 | | | | | | | | | | | | | | | | |
| 4 | ♂AZ3 | CEP192+/- | 58 | N | | | | | | | | | | | | | | 0 | 0 |
| | ♀AX6 | WT | 64 | | | | | | | | | | | | | | | | |
| 5 | ♂BD1 | CEP192+/- | 62 | 30 | 5 | 20 | 2 | 26 | 6 | 39 | 8 | 36 | 10 | 50 | 5 | N | | 6 | 36 |
| | ♀BF5 | WT | 61 | | | | | | | | | | | | | | | | |
| 6 | ♂BD3 | CEP192+/- | 62 | 36 | 9 | N | | | | | | | | | | | | 1 | 9 |
| | ♀CH1 | WT | 58 | | | | | | | | | | | | | | | | |
| 7 | ♂BC1 | CEP192+/- | 61 | 24 | 8 | 25 | 5 | 23 | 9 | 30 | 4 | N | | | | | | 4 | 26 |
| | ♀CH3 | WT | 58 | | | | | | | | | | | | | | | | |
| 8 | ♂BC4 | CEP192+/- | 61 | 24 | 8 | 27 | 7 | 44 | 8 | 35 | 10 | 24 | 9 | 52 | 4 | N | | 6 | 46 |
| | ♀CH9 | WT | 58 | | | | | | | | | | | | | | | | |
| 9 | ♂BB1 | CEP192+/- | 63 | 22 | 6 | 49 | 4 | 30 | 2 | N | | | | | | | | 3 | 12 |
| | ♀BR1 | WT | 60 | | | | | | | | | | | | | | | | |
| 10 | ♂BC1 | CEP192+/- | 62 | 24 | 8 | 25 | 8 | 29 | 5 | 53 | 3 | N | | | | | | 4 | 24 |
| | ♀CH3 | WT | 57 | | | | | | | | | | | | | | | | |

**Supplementary Table 11:** Reproduction of persistent mating wild-type male mice with wild-type females for 210 days. *: days from mate to give the first birth; **: days from giving the first birth to the second birth; N: no birth

| Mouse Pairs | | Genotype | Age (days) | Persistent mating for 210 days | | | | | | | | | | | | | | | |
|---|---|---|---|---|---|---|---|---|---|---|---|---|---|---|---|---|---|---|---|
| | | | | Mating days * | Litter 1 Pups | Mating days ** | Litter 2 Pups | Mating days | Litter 3 Pups | Mating days | Litter 4 Pups | Mating days | Litter 5 Pups | Mating days | Litter 6 Pups | Mating days | Litter 7 Pups | Overall Litters | Overall Pups |
| 1 | ♂AX1 | WT | 59 | 24 | 7 | 21 | 9 | 36 | 12 | 29 | 9 | 22 | 7 | 33 | 9 | 44 | 4 | 7 | 57 |
| | ♀AW1 | WT | 61 | | | | | | | | | | | | | | | | |
| 2 | ♂BJ5 | WT | 59 | 19 | 7 | 24 | 6 | 20 | 4 | 22 | 6 | 23 | 5 | 24 | 5 | 48 | 3 | 7 | 36 |
| | ♀AW5 | WT | 61 | | | | | | | | | | | | | | | | |
| 3 | ♂JJ1 | WT | 59 | 24 | 9 | 36 | 7 | 22 | 9 | 25 | 9 | 24 | 4 | 20 | 5 | 25 | 7 | 7 | 50 |
| | ♀JX2 | WT | 58 | | | | | | | | | | | | | | | | |
| 4 | ♂CX1 | WT | 62 | 38 | 9 | 24 | 7 | 21 | 7 | 27 | 7 | 32 | 5 | 22 | 3 | 28 | 4 | 7 | 42 |
| | ♀JX3 | WT | 58 | | | | | | | | | | | | | | | | |
| 5 | ♂J27 | WT | 60 | 24 | 4 | 38 | 2 | 33 | 11 | 23 | 8 | 23 | 9 | 25 | 3 | 20 | 6 | 7 | 43 |
| | ♀J13 | WT | 60 | | | | | | | | | | | | | | | | |
| 6 | ♂J28 | WT | 60 | 24 | 6 | 20 | 7 | 37 | 7 | 35 | 9 | 22 | 4 | 24 | 3 | 22 | 7 | 7 | 43 |
| | ♀J14 | WT | 60 | | | | | | | | | | | | | | | | |
| 7 | ♂AN1 | WT | 63 | 29 | 8 | 22 | 9 | 39 | 9 | 22 | 6 | 38 | 8 | 24 | 7 | 33 | 8 | 7 | 55 |
| | ♀HC4 | WT | 59 | | | | | | | | | | | | | | | | |
| 8 | ♂HB3 | WT | 61 | 22 | 7 | 44 | 10 | 34 | 8 | 70 | 4 | N | | | | | | 4 | 29 |
| | ♀M6 | WT | 63 | | | | | | | | | | | | | | | | |
| 9 | ♂IM4 | WT | 57 | 21 | 8 | 29 | 7 | 31 | 6 | 50 | 8 | 22 | 9 | 24 | 2 | N | | 6 | 40 |
| | ♀BG4 | WT | 65 | | | | | | | | | | | | | | | | |
| 10 | ♂BG6 | WT | 65 | 20 | 5 | 43 | 8 | 21 | 4 | 23 | 3 | 23 | 4 | 22 | 8 | 24 | 5 | 7 | 37 |
| | ♀BV1 | WT | 57 | | | | | | | | | | | | | | | | |

**Supplementary Table 12:** Original data of the testis-weight and body weight for infertile *Cep192*$^{+/-}$, *Cep192*$^{+/M}$ and *Cep192*$^{+/+}$ mice.

**Note:** Fertile *Cep192*$^{+/-}$, *Cep192*$^{+/M}$ mice showed normal or nearing-to-normal testis size. In here, we presented only the testis size data for *Cep192*$^{+/-}$, *Cep192*$^{+/M}$ mice with infertility.

| Mice ID | Genotype | Body Weight (g) | Testis ID | Testis Weight (mg) | Testis Weight/ Body Weight (mg/g) |
|---|---|---|---|---|---|
| WT1 | *Cep192* $^{+/+}$ | 28.79 | Testis 1 | 107.30 | 3.73 |
| | | | Testis 2 | 95.80 | 3.33 |
| WT2 | | 27.02 | Testis 3 | 103.20 | 3.82 |
| | | | Testis 4 | 97.60 | 3.61 |
| WT3 | | 26.97 | Testis 5 | 103.00 | 3.82 |
| | | | Testis 6 | 103.00 | 3.82 |
| WT4 | | 26.90 | Testis 7 | 101.20 | 3.76 |
| | | | Testis 8 | 102.50 | 3.81 |
| WT5 | | 27.57 | Testis 9 | 106.60 | 3.87 |
| | | | Testis 10 | 105.30 | 3.82 |
| WT6 | | 27.95 | Testis 11 | 101.20 | 3.62 |
| | | | Testis 12 | 102.70 | 3.67 |
| HET1 | *Cep192* $^{+/-}$ | 27.42 | Testis 1 | 75.20 | 2.74 |
| | | | Testis 2 | 88.60 | 3.23 |
| HET2 | | 26.16 | Testis 3 | 65.90 | 2.52 |
| | | | Testis 4 | 70.00 | 2.68 |
| HET3 | | 32.60 | Testis 5 | 80.50 | 2.47 |
| | | | Testis 6 | 80.90 | 2.48 |
| HET4 | | 31.50 | Testis 7 | 71.20 | 2.26 |
| | | | Testis 8 | 72.50 | 2.30 |
| HET5 | | 27.43 | Testis 9 | 27.80 | 1.01 |
| | | | Testis 10 | 25.50 | 0.93 |
| HET6 | | 26.90 | Testis 11 | 33.50 | 1.25 |
| | | | Testis 12 | 32.40 | 1.20 |
| HET1 | *Cep192* $^{+/M}$ | 27.83 | Testis 1 | 65.50 | 2.35 |
| | | | Testis 2 | 75.70 | 2.72 |
| HET2 | | 28.47 | Testis 3 | 80.40 | 2.82 |
| | | | Testis 4 | 80.90 | 2.84 |
| HET3 | | 29.87 | Testis 5 | 94.50 | 3.16 |
| | | | Testis 6 | 95.70 | 3.20 |
| HET4 | | 27.62 | Testis 7 | 79.00 | 2.86 |
| | | | Testis 8 | 83.02 | 3.01 |
| HET5 | | 27.18 | Testis 9 | 54.02 | 1.99 |
| | | | Testis 10 | 61.34 | 2.26 |
| HET6 | | 29.25 | Testis 11 | 70.50 | 2.41 |
| | | | Testis 12 | 69.80 | 2.39 |

**Supplementary Table 13:** Primers synthesized for the Minigene Assay

| Name | Primer (from 5' to 3') |
|---|---|
| 40356-CEP192-F | gccatatgtcaggtgcccag |
| 40677-CEP192-F | attaatggaattatagtaag |
| 42479-CEP192-R | aacatacaataagttgaatt |
| 42726-CEP192-R | agtctgtggaataaggacca |
| MINI-CEP192-KpnI-F | ggtaggtaccgcagaccttccagaaacttt |
| MINI-CEP192-BamHI-R | tagtggatcctcatcctaaataataaatta |
| CEP192-MUT-F | aaaaagaaatggcttatcttaaccatgatc |
| CEP192-MUT-R | gatcatggttaagataagccatttcttttt |

**Supplementary Figure 10:** The plasmid construction for the Mini gene Assay

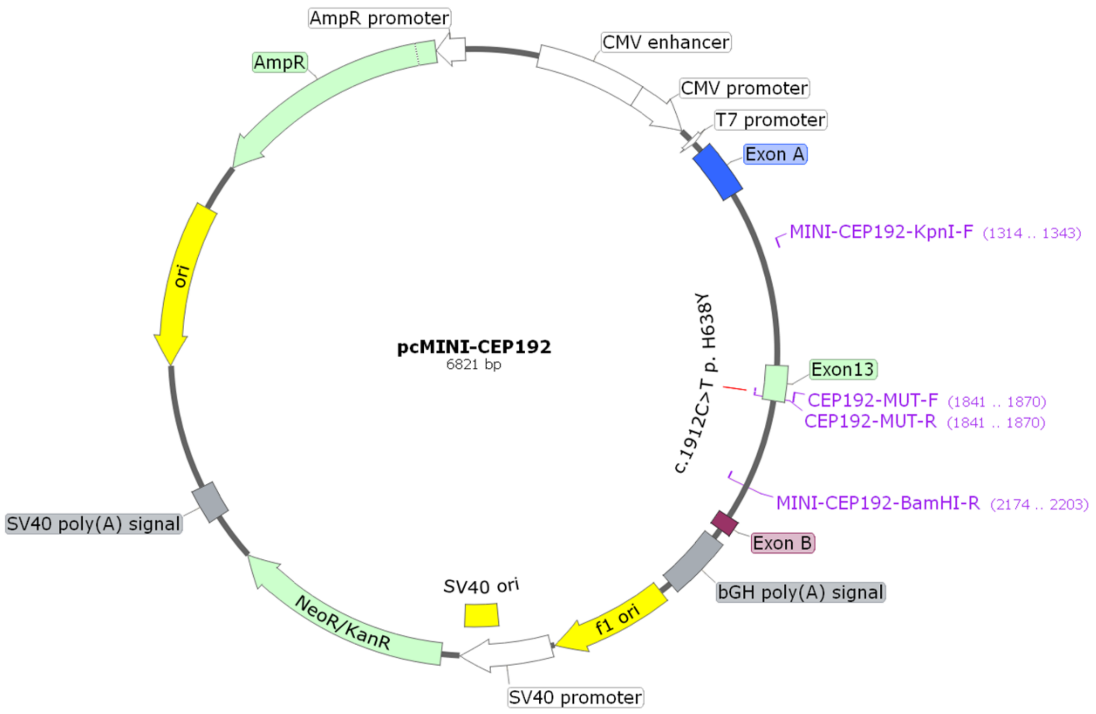

**Supplementary Table 14:** Results of two rounds of karyotyping for patient B

| **FIRST ROUND** | | **SECOND ROUND** | |
|---|---|---|---|
| **Karyotype** | **Count** | **Karyotype** | **Count** |
| 92,XXXX (*) | 7 | 92, XXXX | 29 |
| 47,XXX | 1 | 45,X0 | 1 |
| 47,XX,+mar | 1 | 47,XX,+mar | 2 |
| 46,XX,del(3)(q27) | 1 | 48,XX,+1,+11 | 1 |
| 46,XX,del(6)(p21) | 1 | 45,XX,-5 | 1 |
| 48,XX,+8,+18 | 1 | 46,XX,del(5)(p15) | 1 |
| 45,XX,-13 | 1 | 45,XX,-6 | 1 |
| 45,XX,-14 | 1 | 45,XX,-7 | 1 |
| 47,XX,+15 | 1 | 45, XX,-8, t(2;11)(p10; q12) | 1 |
| 45,XX,-16 | 1 | 48,XX,+8,+10 | 1 |
| 45,XX,-18 | 2 | 46,XX, del(7)(q32) | 1 |
| 47,XX,+18, del (6)(q22) | 1 | 47,XX,-8, +mar1, +mar2 | 1 |
| 47,XX,+20 | 1 | 45,XX,-9 | 1 |
| 46,XX,del(20)(q13) | 1 | 48,XX,+11,+22 | 1 |
| 46,XX,del(21)(q22) | 1 | 46,XX,del(16)(q21) | 1 |
| 45,XX,-22 | 1 | 45,XX,-12 | 2 |
| 46,XX | 58 | 45,XX,-14 | 1 |
| | | 45,XX, -16 | 1 |
| | | 47, XX,+16 | 1 |
| | | 47,XX,+18 | 4 |
| | | 45,XX,-18 | 2 |
| | | 46,XX,del(18)(q22) | 1 |
| | | 46, XXX,-19 | 1 |
| | | 47, XX,+19 | 1 |
| | | 45,XX,-20 | 1 |
| | | 45,XX,-22 | 2 |
| | | 45, XX, -22, del (16)(q22) | 1 |
| | | 46, XX | 59 |
| 81 metaphase cells analyzed | | 121 metaphase cells analyzed | |
| 23 abnormal cells, 28.40% | | 62 abnormal cells, 41.06% | |

*in tetraploidy cells, still several of them existed anuploidy, such as 91, XXXX, -11, which were not shown cell by cell in here.

**Supplementary Table 15:** Result of karyotyping for patient A

| **Karyotype** | **Count** |
|---|---|
| 92,XXYY | 21 |
| 46,XY,del(2)(p23), del(16)(q23) | 1 |
| 45,XY,-4 | 2 |
| 46, XY, del(5)(p15) | 1 |
| 45, XY, -6 | 1 |
| 46, XY, -13, +18 | 1 |
| 46, XY, del (16)(q23) | 2 |
| 47,XY,+18 | 6 |
| 46,XY,+18, -21 | 1 |
| 46, X, -Y, +18 | 2 |
| 45, XY, -19 | 2 |
| 45, XY, -21 | 4 |
| 46, XY | 43 |
| 87 metaphase cells analyzed | |
| 44 abnormal cells, 51.16% | |

**Supplementary Table 16:** Results of two rounds of karyotyping for patient C

| **FIRST ROUND** | | **SECOND ROUND** | |
|---|---|---|---|
| **Karyotype** | **Count** | **Karyotype** | **Count** |
| 92,XXYY | 2 | 92,XXYY | 3 |
| 46,XY,-2,+MAR | 1 | 45,XY,-2 | 1 |
| 46,XY,t(2;5)(q12;q13) | 1 | 45,XY,-4 | 1 |
| 45,XY,-15 | 1 | 46,XY,-4,+RING | 1 |
| 46,X,+18,-Y | 1 | 45,XY,-5 | 1 |
| 45,XY,-19 | 1 | 45,XY,-6 | 1 |
| 45,XY,-20 | 1 | 45,XY,-8 | 1 |
| 45,XY,-22 | 1 | 45,XY,-9 | 1 |
| 46,XY | 48 | 45,XY,-14 | 1 |
| | | 45,XY,-15 | 1 |
| | | 45,XY,-17 | 1 |
| | | 45,XY,-22 | 1 |
| | | 45,X | 3 |
| | | 46,XY | 89 |
| 57 metaphase cells analyzed | | 106 metaphase cells analyzed | |
| 9 abnormal cells, 15.79% | | 17 abnormal cells, 16.04% | |

**Supplementary Table 17:** Result of karyotyping for patient D

| Karyotype | Count |
|---|---|
| 92, XXYY | 2 |
| 47, XY, +8 | 1 |
| 45, XY, -7 | 1 |
| 45, XY, -10 | 1 |
| 45, XY, -15 | 1 |
| 45, XY, -18 | 1 |
| 45, XY, -19 | 2 |
| 45, XY, -21 | 1 |
| 46, XY | 54 |
| 64 metaphase cells analyzed | |
| 10 abnormal cells, 15.63% | |

**Supplementary Table 18:** Result of karyotyping for patient E

| **Karyotype** | **Count** |
|---|---|
| 90,XXYY (Nearing to tetraploidy) | 1 |
| 59,XY,+5,+6,+8,+8,+9,+10,+13,+13,+14,+15,+17,+17,-18,+19,+20 | 1 |
| 48,XY,+10,+12 | 1 |
| 48,XY,+9,+13 | 1 |
| 48,XY,+2mar | 1 |
| 47,XYY | 2 |
| 47,XY,+mar | 1 |
| 46,XY,t(7;14)(q36;q13) | 1 |
| 45,X | 3 |
| 45,XY,-3 | 1 |
| 45,XY,-4 | 1 |
| 45,XY,-9 | 1 |
| 45,XY,-11 | 1 |
| 45,XY,-12 | 2 |
| 45,XY,-13 | 3 |
| 45,XY,-14 | 2 |
| 45,XY,-15 | 2 |
| 45,XY,-16 | 1 |
| 45,XY,-18 | 2 |
| 45,XY,-19 | 1 |
| 45,XY,-21 | 2 |
| 45,XY,-22 | 3 |
| 44,XY,-20,-20 | 1 |
| 46,XY | 251 |
| 286 metaphase cells analyzed | |
| 35 abnormal cells, 12.24% | |

**Supplementary Table 19:** Result of karyotyping for patient G

| **T00634 at 29 years old** | |
|---|---|
| **Karyotype** | **Count** |
| 92,XXYY | 1 |
| 60,XXY,+1,+1,+2,+2,+3,+3,+4,+5,+6,+6, +7,+10, +11,+12,-13,-18,+21 | 1 |
| 50,XY,+1,+2,+3,+5,+13,-14,-19,+22 | 1 |
| 47,XY,+14 | 1 |
| 44,XY,-9,-9 | 1 |
| 45,XY,-7 | 1 |
| 45,XY,-12 | 2 |
| 45,XY,-13 | 1 |
| 45,XY,-15 | 1 |
| 45,XY,-18 | 1 |
| 45,XY,-21 | 1 |
| 45,XY,-22 | 1 |
| 46,XY | 64 |
| 77 metaphase cells analyzed | |
| 13 abnormal cells, 21.15% | |
